# Cognitive Impairment in Paediatric PCNS: A Systematic Review and Meta-analysis

**DOI:** 10.1101/2025.05.29.25328543

**Authors:** Lauren Falcone, Nina Riddell, Melanie J. Murphy

## Abstract

Post-acute sequalae of Coronavirus disease-19 (COVID-19 [PASC]) are defined as the persistence of existing or new symptoms for a period extending beyond initial COVID-19 infection. Post COVID Neurological Syndrome (PCNS) relates to the persistent cognitive and neurological deficits characteristic of PASC. This includes significant changes in attention and memory function in adult and geriatric populations, with such impairments notably impacting quality of life. However, despite reports of similar cognitive changes in paediatric patients, this issue is yet to be systematically investigated. This systematic review and meta-analysis synthesised literature reporting on the prevalence of clinically significant cognitive impairment in paediatric PASC using DSM-5 cognitive domains to categorise study outcomes. Final literature searching was completed on 15^th^ of November, 2024 across four databases combining the following keywords: ‘COVID-19’, ‘cognition’ and ‘paediatric’. Included studies examined standardised psychometric or parent report measures of cognition in children and adolescents with a PASC diagnosis. Studies were excluded if participants had prior cognitive impairments or comorbidities. Risk of bias was assessed using Joanna Briggs Institute Critical Appraisal Tool’s Checklist for Analytical Cross-sectional Studies. Results revealed that between 35-55% of paediatric PASC patients were ‘at risk of impairment’ or showed ‘clinically significant’ impairment in complex attention, learning and memory, working memory and executive function. Further research is needed to assess impacts of infection severity and repeated infection. However, this meta-analysis provides insights into the nature of PCNS-associated complications to aid more detailed management strategies for children and adolescents.

---

As society moves into a post-pandemic world, the Coronavirus Disease 2019 (COVID-19) pandemic continues to be one of the most influential global events with lasting impacts felt worldwide (Chen et al., 2022; Sudre et al., 2021). Australia accounts for 11.9 million of the 776 million confirmed COVID-19 cases worldwide, though the true infection rate is likely higher due to underreporting, particularly in children and adolescents (Levine, 2022; World Health Organisation, 2024). As COVID-19 begins to approach endemic status, it is estimated that over 71% of the Australian population have been infected with the virus as of 2022 (Australian COVID-19 Serosurveillance Network, 2023; Townsend et al., 2023). Although the average COVID-19 infection period typically lasts for 14 days, individuals often experience and report persistent symptoms, with approximately 20% of adults diagnosed with COVID-19 living with ongoing health conditions as a result (Bull-Otterson et al., 2022; Ceban et al., 2022; Di Toro et al., 2021).

This experience of ongoing health impacts can result in the diagnosis of post-acute sequelae of COVID-19 (PASC), defined as the persistence of new or existing symptoms at least three months after infection (Bonilla et al., 2023; World Health Organisation, 2022). The global prevalence of PASC ranges between 10-30%, with varying levels of severity and symptom longevity (Ceban et al., 2022; Levine, 2022; Wang et al., 2023). It is estimated that one third of PASC patients do not fully recover from their symptoms and still experience at least one symptom after a mean of 125 days from onset (Alwan & Johnson, 2021; Petersen et al., 2021; Seessle et al., 2022). PASC, often used interchangeably with long COVID or Post-COVID Condition (PCC), can encompass a wide variety of symptoms, including respiratory symptoms, loss of sense of smell, mental fatigue, ‘brain fog’ and chronic migraines (Ceban et al., 2022; Chen et al., 2022; Di Toro et al., 2021; Masserini et al., 2025; Sudre et al., 2021; Wijeratne & Crewther, 2020; World Health Organisation, 2023).

The high incidence of ongoing neurological symptoms and cognitive impairment following COVID infection is termed Post-COVID Neurological Syndrome (PCNS) (Wijeratne & Crewther, 2020), with the PCNS profile consistent with the core symptom cluster recommended for screening in cases of suspected PCC (Masserini et al., 2025). The manifestation of PCNS is not an unexpected consequence of the multi-system impact of COVID infection, with emerging evidence highlighting significant alteration to immunological (Menezes et al., 2024) and neurological, cognitive and psychological function (Cerioli et al., 2024; Joshi et al., 2024; Masserini et al., 2025; Narayanan et al., 2025) following infection. Vaccination may not be a protective factor against development of PCNS (Mukherjee et al., 2025), and interestingly, the risk of PCNS appears to differ between ethnicities (Zhang et al., 2025), genders and across age groups (Gonzalez Aleman et al., 2025; Su et al., 2024), with younger and middle-aged adults showing higher incidence and impact of PCNS on cognitive dysfunction and quality of life measures (Choudhury et al., 2025).

Although PCNS imposes a significant quality of life burden for adults due to ongoing health, lifestyle, and socio-economic impacts (Alwan & Johnson, 2021; Petersen et al., 2021), the detrimental effect of PCNS on the paediatric population is less well characterised. This has been in part due to greater focus on other physical symptoms, a lack of adequate screening tools or protocols, or difficulties in interpreting patient self-report or parent report (Masserini et al., 2025; Patel et al., 2025; Pour Mohammadi et al., 2024; Tso et al., 2024). The impact of PCNS on the cognitive abilities of children is of particular concern as optimal cognitive function is imperative to cognitive development, especially in two of the most influential yet fragile developmental periods, childhood and adolescence (Arain et al., 2013; Jirout et al., 2019).

## Post-COVID Neurological Syndrome Symptomology & Pathophysiology

In both adult and paediatric populations, the cognitive impairment component of PCNS often presents alongside other psychological and physical PASC symptoms (Narayanan et al., 2025; World Health Organisation, 2023; Wulf Hanson et al., 2022). Evidence suggests that these symptoms can worsen over time (Davis et al., 2021; Jason et al., 2021; Sekendiz et al., 2024).

Mental health disorders such as anxiety and depression are often associated with these symptoms because of the inter-relationship between immune system inflammation and neural function (Lamontagne et al., 2021; Narayanan et al., 2025), and due to the symptom burden on an individual’s daily functioning (Luedke et al., 2024).

Symptom manifestations of PASC are like COVID-19 in that they are clinically heterogeneous and involve multiple organs and bodily systems (Di Toro et al., 2021; Masserini et al., 2025). This can be seen across respiratory, cardiovascular, digestive, and muscular systems (Di Toro et al., 2021; Li et al., 2023). Multi-organ involvement in PASC can be observed throughout the heart, liver, and brain (Li et al., 2023). Cognitive change in PASC is highly correlated with increased inflammatory blood biomarkers as well as cortical area changes and hippocampal and grey matter volume shrinkages (Invernizzi et al., 2023; Kwan et al., 2024; Lai et al., 2023; Rothstein, 2023). Given these changes in cognitive function, it is imperative to develop a framework through which to examine these impairments more closely.

## Conceptualising Cognition

When conceptualising cognitive function to assess specific areas of impairment, cognition can be broken down into six domains, as per the Diagnostic and Statistical Manual of Mental Disorders (DSM-5; (American Psychiatric Association, 2013): complex attention, learning & memory, language, executive function, perceptual motor-function, and social cognition (Sachdev et al., 2014).

Complex attention refers to the ability to control, shift and divide attentional focus across multiple stimuli (Daffner et al., 2015). Learning and memory encompasses acquisition and storage of new information and can also include implicit learning, or the ability to learn without intention or conscious effort (Squire, 1987; Vinter et al., 2010). Language can include verbal comprehension skills such as word finding or fluency and can also include the ability to utilise grammar and syntax (Harvey, 2019; Sachdev et al., 2014). Executive function is a broad cognitive domain that notably includes working memory, that is, the process where information is stored temporarily for accessibility or manipulation in cognitive tasks (Harvey, 2019).

Executive function also involves higher order thinking processes like planning, inhibition, and problem solving (Sachdev et al., 2014). Perceptual-motor function encompasses the ability to combine spatial awareness with motor function, and is responsible for perceptual-motor coordination, a process utilised in handwriting (Tseng & Chow, 2000). Social cognition involves the ability to recognise and interact with socially relevant information, and is responsible for how an individual thinks, feels, and behaves in social situations (Adolphs, 2001).

There are many theoretical models of cognition that posit cognitive function as a hierarchical structure (Dams-O’Connor & Gordon, 2013; Fisher, 2019; Harvey, 2019). Within these models, sensory, attentional and perceptual cognitive domains are considered to have the greatest impact on overall cognition, and when their function is compromised, the impacts can be observed across other domains (Harvey, 2019). Complex attention can be considered as one of the core foundational domains, notably in children and adolescents due to the further developmental impacts (Hendry et al., 2016; Rothbart et al., 2011; Ruff & Rothbart, 2001). If an individual is unable to attend and experiences impairments in complex attention, they are likely to also experience impairments across learning and memory function, given that attention directly impacts the ability to encode and store information (Oberauer, 2019; Siegel & Castel, 2018). If a child is unable to learn and remember because of a complex attention dysfunction, lasting impacts can be seen across their developmental lifespan (Hendry et al., 2016).

## Cognitive Impairment in Adults with PCNS

A growing body of evidence suggests that cognition is impaired across multiple domains in adults with PASC. At a broader level of cognition, a meta-analysis conducted by Ceban et al. (2022) found that over a fifth of patients displayed overall cognitive dysfunction at 12 weeks post-COVID-19 infection. In a meta-analysis of 42 studies across four different follow-up periods for infection (30, 60, 90, and 120 days), concentration difficulties were the third most common symptom observed throughout the sample after chronic fatigue and memory impairment (Chen et al., 2022). Interestingly, both Ceban et al. (2022) and (Chen et al., 2022) reported that the degree of impairment was consistent across hospitalised and non-hospitalised individuals, suggesting that development of cognitive issues associated with PCNS is not linearly predictable based on the severity of the initial COVID-19 infection and other risk factors may be involved. Across 66 studies, Fanshawe et al. (2024) meta-analysed cognitive subdomain scores from hospital administered psychometric assessments and found that all domains, but most notably, complex attention, working memory, learning and memory and executive function, were negatively impacted in adults with PCNS when compared to healthy controls.

More recent reports exploring the subject experience of brain-fog and objective assessment of cognitive function in PCC reported that attention and episodic memory disturbances were the most frequent impairments reported (Delgado-Alonso et al., 2025). Interestingly, the degree of cognitive impairment reported by patients was significantly mediated by levels fatigue and depression, and to some extent anxiety. A further study similarly found decline in overall cognitive function, with specific impairments in attention, reaction times, short-term and working memory and mental flexibility (a component of executive function), which were impacted by fatigue, depression and anxiety (Charles James et al., 2025).

While the above analyses provide evidence for cognitive impairments in the PASC population, they do not consider how such impairment may differ across the lifespan. This is problematic when the age-related differences in PCNS symptom presentation described above are considered. Closer examination of findings from adult studies reveals that younger adults (18 to 29 years of age) present with higher prevalences of attention and executive functioning deficits when compared to older adults, suggesting that younger patients may experience greater impacts on cognition following PASC (Davis et al., 2021).

## PCNS in Paediatric Cohorts

Literature published near the beginning of the pandemic suggests that at least 10 percent of all children who test positive for COVID-19 will go on to develop PASC (Di Toro et al., 2021). However, more recent literature has suggested this number may be higher than once anticipated, as COVID-19 cases spike in children due to an overall lower vaccination rate compared to adults (“Long COVID and kids: more research is urgently needed,” 2022; Zheng et al., 2023).

PCNS can impact a child or adolescent’s functional behaviour, and engagement in academic and social settings (Gonzalez-Aumatell et al., 2022). Further, developmental regression is now being reported in children affected by PCNS (Ashkenazi-Hoffnung et al., 2021). Given the developmental importance of cognitive skill acquisition during childhood and adolescence, understanding the impact of PCNS on cognition in children is imperative, particularly in light of evidence showing enduring, and potentially worsening PCNS symptomology without intervention (Arain et al., 2013; Ashkenazi-Hoffnung et al., 2021; Jason et al., 2021).

## Cognitive Impairment in Paediatric PCNS

Examination of specific cognitive impairments in children with PASC frequently identifies difficulties with sustained attention and memory (Gonzalez-Aumatell et al., 2022; Luedke et al., 2024; Scarselli et al., 2023). Both paediatric self-report and parent-report survey-based studies show evidence of several cognitive difficulties. Brackel et al. (2021) revealed that half of the children surveyed displayed concentration difficulties, and 13% exhibited memory loss of some kind. Similarly, confusion and lack of concentration was reported in approximately 10% of paediatric PCNS patients, rising to 11.8% at four month follow up (Buonsenso et al., 2021). Similarly, single cohort, clinic-based studies have found that 30% of children and adolescents experience cognitive disturbances and at 6-month follow-up, this only dropped to 20% (Alghamdi et al., 2022; Weakley et al., 2023). Further, COVID-19 symptom severity has been linked to more severe cognitive difficulties (Avittan & Kustovs, 2023), again raising the question of whether severity of COVID-19 infection may be predictive of future PCNS-related cognitive dysfunction.

Patient and parent observation of cognitive issues have been supported by investigations using standardised hospital-based and neuropsychological assessment tools administered in formal settings. Cognitive assessments, such as the Montreal Cognitive Assessment (MoCa; (Nasreddine et al., 2005) and the Neuropsychological Assessment - Second Edition (NEPSY-II; (Korkman et al., 2007b), examine competence across several cognitive domains and are administered by trained health professionals. In a sample of children aged 8 to 17 affected by PASC assessed using the NEPSY-II, 54% were shown to have impairments in attention and executive function, and 40% displayed memory impairments (Scarselli et al., 2023). Gonzalez-Aumatell et al. (2022) assessed cognitive function through designing the Pediatric Symptom Checklist (PSC) screening tool and identified cognitive dysfunction in 60% of children attending a long-COVID unit. These children were referred for further evaluation by a clinician and of this sample, 63% had sustained attention dysfunction, approximately 50% had impaired executive function, and 30% displayed poor processing speed. Together these findings demonstrate that paediatric PCNS patients and their caregivers perceive the degree of cognitive impairment arising from the condition, and that this is measurable in terms of clinical impact.

Together the findings examined above illustrate that children and adolescents experience cognitive impairment associated with PCNS. Of note is that children themselves report noticing an impact on their functioning (Brackel et al., 2021; Sterky et al., 2021). However, despite WHO’s (2023) observation that PCNS can impact a child’s ability to achieve and maintain developmental milestones, and the notable focus on cognitive impairment in adult populations (Rumain et al., 2021), a systematic synthesis of evidence regarding the exact prevalence and nature of cognitive impairment associated with PCNS for paediatric populations is lacking. This disruption to developmental milestones can manifest in further disruption of education that can be quantified through absenteeism and low involvement in extracurricular activities, both of which have further implications on a child’s socialisation (Miller et al., 2024). Developing a more complete understanding of this is important to characterise the cognitive risk profile of paediatric patients affected by PCNS.

Thus, we aimed to systematically review and meta-analyse literature reporting the prevalence of cognitive impairments associated with PCNS in paediatric populations while following standardised Preferred Reporting Items for Systematic Reviews and Meta-Analyses (PRISMA; (Page et al., 2021) protocols. We achieved this through database search strategies targeting studies of PASC (and PASC synonyms) examining cognition in child and adolescent populations. Where possible we aimed to use the DSM-5 (American Psychiatric Association, 2013) cognitive domains, such as complex attention, learning & memory, and executive function to more closely explore the specific profile of impairment in children with PASC, and to gain overall a better understanding of the complications children and adolescents may experience, promoting more targeted symptom management and support.

## Methods

This systematic review and meta-analysis was conducted in line with the PRISMA 2020 guidelines (Page et al., 2021) and was prospectively registered with Open Science Framework (OSF) on the 5th of May 2024 (registration DOI: 10.17605/OSF.IO/7SC4M).

### Search Strategy

Final literature searches were conducted on the 15^th^ of November 2024 across PubMED, Ovid Medline, Ovid Embase and Web of Science databases to achieve maximum literature recall for studies investigating cognitive impairment in children post-COVID-19 (Bramer et al., 2017). The search strategy, which was reviewed by a senior librarian, was adapted according to each database’s keyword and subject heading parameters and is summarised in Table 1.

**Table 1.** Search Strategy.

| Row | Search Command |
| --- | --- |
| 1 | Coronavirus disease 2019/ |
| 2 | COVID-19.mp. |
| 3 | Severe acute respiratory syndrome coronavirus 2/ |
| 4 | SARS-CoV-2.mp. |
| 5 | Post-acute sequelae of COVID-19.mp. |
| 6 | PASC.mp. |
| 7 | Long covid.mp. |
| 8 | 1 or 2 or 3 or 4 or 5 or 6 or 7 |
| 9 | Cognit*.mp. |
| 10 | Neuropsych*.mp. |
| 11 | 9 or 10 |
| 12 | Child*.mp. |
| 13 | P?ediatric.mp. |
| 14 | Adolescen*.mp. |
| 15 | Teen*.mp. |
| 16 | 12 or 13 or 14 or 15 |
| 17 | 8 and 11 and 16 |
| 18 | Limit 17 to (English language and yr="2020 -Current") |

Note. The symbol (\*) allows for truncation searching while the symbol (?) allows for the search strategy to capture British and American spelling variations. The term (.mp.) allows for a search across an article's title, abstract, and subject headings. The forward slash (/) allows for a search across only subject headings.

### Selection Criteria

Search results from each database were uploaded into Covidence (Veritas Health Innovation, 2023) for screening. After removal of duplicates, two independent reviewers conducted title and abstract and full text screening on all articles against the criteria outlined below. Conflicts related to the inclusion or exclusion of studies were resolved through consensus discussion between three reviewers.

Full text exclusion criteria are summarised in Table 2. Studies selected for inclusion in the analysis were original, peer-reviewed studies published in the English language after January 2020. There was no limitation on recruitment setting. Participants in each study were required to be under 18 years of age. Given the recency of PASC literature and to account for diagnostic criteria updates, PASC groups needed to adhere to the diagnostic criteria of PASC as stated by their country’s primary health organisation at the time of experiment (i.e., WHO, CDC). Studies could include participants who were or were not hospitalised due to their COVID-19 infection. Studies could compare PASC groups with comparators (e.g., healthy controls, COVID-recovered). Included studies assessed cognitive function through validated psychometric tests or parent- and self-report measures. In light of variability between the minimum age guidelines across several psychometric tests, self-report, and parent report measures, no minimum age limit was imposed to ensure capture of measures normed for early childhood (Achenbach, 1991; Korkman et al., 2007a; Roman et al., 2014).

**Table 2.** Full Text Exclusion Criteria.

| Full Text Exclusion Criteria |
| --- |
| 1. Full text not available |
| 2. Full text not in English |
| 3. Adult population (18+) |
| 4. Experimental group does not meet PASC criteria |
| 5. Pre-existing diagnosis that may impact cognition |
| 6. No objective or validated measure of cognition |
| 7. Testing time post-infection unspecified or unclear |
| 8. Appropriate quantitative data not reported |

Studies were excluded if participants had a prior diagnosis involving cognitive impairment or any other medically diagnosed condition that could impact cognitive function irrespective of COVID-19 infection status. In cases where the follow-up time post-infection was unclear or not specified, these studies were also excluded as it was not possible to determine whether participants met accepted PASC diagnostic criteria. Studies were also excluded if appropriate quantitative data was not reported and was unable to be retrieved after contact with authors.

### Data Extraction and Synthesis

Data from studies that progressed through full text screening were manually extracted. As shown in Table 3, publication details, participant characteristics, outcome measures and outcome variables were extracted in preparation for final analyses. Group characteristics that could impact the degree of impairment observed including hospitalisation status and symptom severity were extracted for potential subgroup analyses.

**Table 3.**
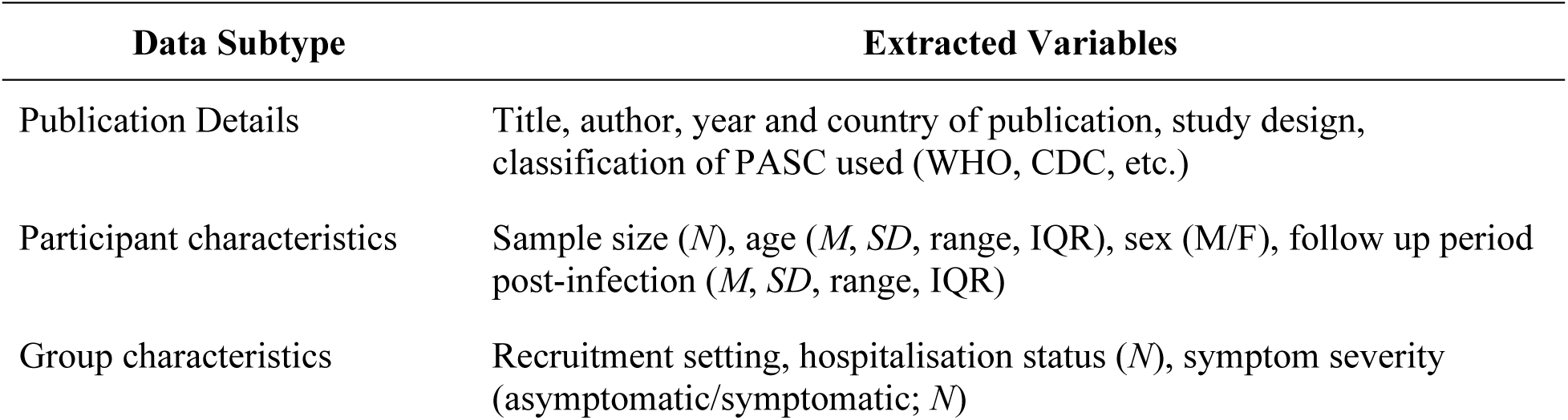
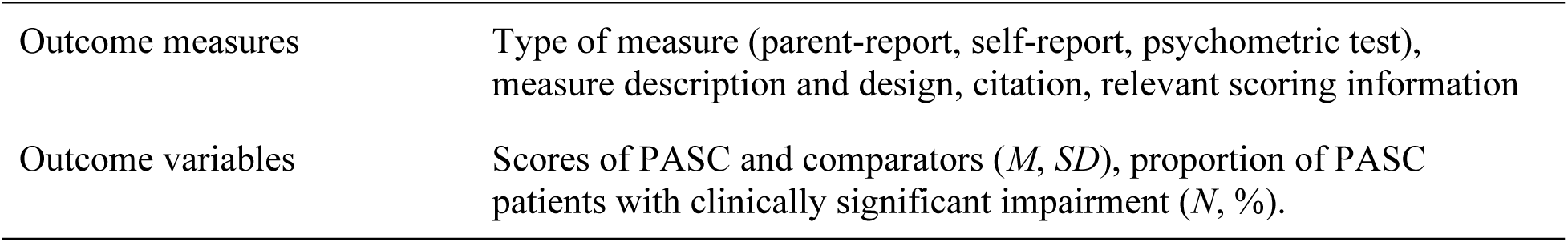
Data Extraction Strategy for Studies Included in the Analysis.

### Quality Assessment

Quality and risk of bias assessment was conducted using the *Joanna Briggs Institute (JBI) Critical Appraisal Tool’s* Checklist for Analytical Cross-Sectional Studies (Moola et al., 2017) based on recommendations from the Australian National Health and Medical Research Centre (2019). Quality assessment was conducted by two independent reviewers who evaluated whether 1) the inclusion criteria for the sample was explicitly defined, 2) the study subjects and settings were described in sufficient detail, 3) the exposure was measured in a valid and reliable manner, 4) objective, standard criteria was used for measurement of conditions, 5) confound variables were identified, 6) there were strategies in place for the management of these confounds, 7) the outcomes were measures in a valid and reliable manner and 8) the statistical analysis was appropriate. Items were rated either ‘Yes’ (criterion met indicating a low risk of bias), ‘No’ (criterion not met indicating a high risk of bias), ‘Unclear’ (insufficient information to meet or not meet criteria) or ‘Not Applicable’ (criterion not relevant).

### Statistical Analysis

Statistical analysis was conducted in Jamovi (Version 2.3.21) using the Meta-Analysis for Jamovi module (MAJOR, Version 1.2.4). A meta-analysis of prevalence was performed on eligible studies to assess the pooled prevalence of cognitive impairment across studies. Arcsine square root transformed proportion analysis was used to stabilise variance across the data and control for publication bias (G. Rucker et al., 2008). Heterogeneity was assessed through the *I*^2^ statistic with 95% confidence intervals. Values ranging between 0-40% constitute minimal heterogeneity while values ranging between 30-60%, 50-90% and 75-100%, represent moderate, substantial, and considerable heterogeneity, respectively (Higgins et al., 2019). The tau squared (*τ* ^2^) statistic was also used to assess heterogeneity as recommended by (G. Rucker et al., 2008).

Funnel plots and the *Q* statistic are not deemed informative for meta-analyses with less than ten data points and thus were not examined in the current analysis (Begg & Mazumdar, 1994). Publication bias was assessed through interpretation of the rank correlation test for funnel plot asymmetry as recommended by Rucker et al. (2011) and the *Cochrane Handbook for Systematic Reviews of Interventions* (Page et al., 2024). Kendall’s tau statistic was interpreted based on statistical significance, where *p* < .05 indicates funnel plot asymmetry that can be attributed to publication bias (Gjerdevik & Heuch, 2014).

Sensitivity analysis was conducted to aid in controlling for heterogeneity and minimise the effects of influential cases where multiple data points from the same group of participants was represented in the analysis (Higgins et al., 2019; Migliavaca et al., 2022). This was completed to evaluate the robustness and reliability of any findings from prevalence analyses based on the inclusion or exclusion of certain data points and examine how these change in line with different measurement tools used across different studies (Deeks et al., 2023).

In complement of the prevalence analyses, narrative synthesis was presented for studies found to be unviable for meta-analysis. To aid interpretation of outcomes from these studies, Hedges’ *g* effect size was calculated to examine the magnitude of effects between PASC groups and comparison groups reported (Hedges, 1981). Effect sizes were interpreted based on Cohen’s classifications where coefficients of 0.2, 0.5 and 0.7 respectively represent small, medium and large effects (Cohen, 1988).

## Results

Final database searching retrieved 7647 studies meeting the search criteria. As indicated in the PRISMA chart in Figure 1 below, 3375 duplicate studies were removed, and 4271 title and abstracts were screened according to the selection criteria. Sixty-six studies progressed into the final full text screening stage. The most common reason for full text exclusion was samples that included adult participants, where 17 studies were excluded on this basis. A total of seven studies were selected for inclusion into the systematic review and meta-analysis. A summary of the extracted studies’ characteristics is presented in Table 4.

**Figure 1.**
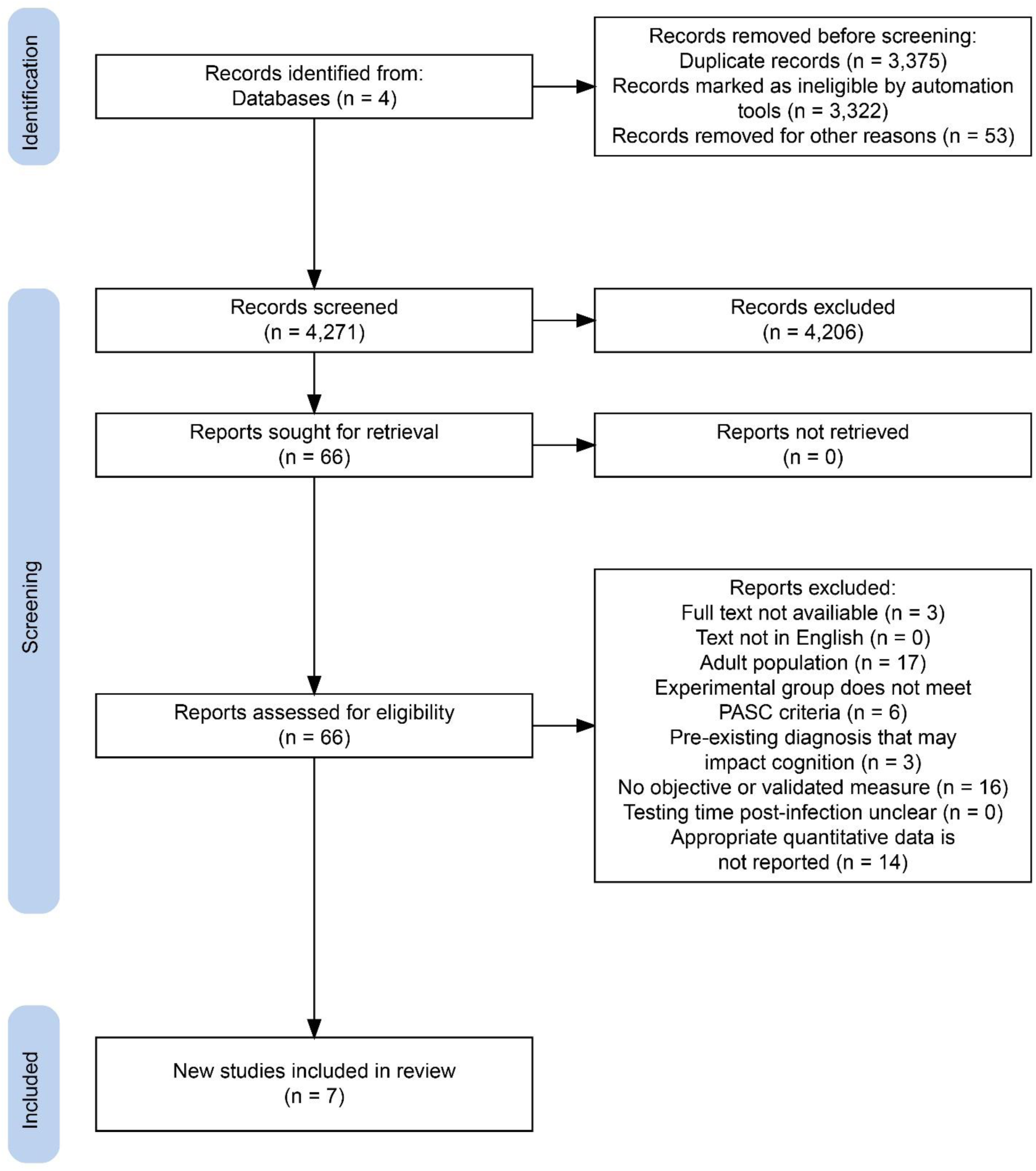
PRISMA Flowchart of Study Screening and Selection Process.

**Table 4.** Summary Characteristics of Included Studies.

| Study Name | Country<br>(PASC<br>definition<br>used) | Study/<br>Recruit-<br>ment<br>Setting | Follow up time<br>post-COVID-19<br>infection,<br>days,<br>*M(SD)* | Sample Size<br>*N* (F/M) | *n* of<br>hospitalised<br>and non-hospitalised<br>participants | Age, years<br>*M (SD)* | Outcome<br>Measures | Outcome<br>Variables |
| --- | --- | --- | --- | --- | --- | --- | --- | --- |
| Ertesvag et al. (2023) | Norway<br>(WHO) | COVID<br>patient<br>population | 245<br>(205-254)*** | 9 (N/A) | N/A | 13.2 (1.7) | CFS* | *n* of the sample<br>reported meeting<br>criteria for<br>significant<br>impairment |
| Guido et al. (2022) | Italy<br>(WHO) | PASC<br>clinic | 31 (N/A) | 322 (165/167) | N/A | 9.53 (3.73) | CBCL* | *n* of sample scoring<br>in the borderline/<br>clinical range,<br>indicating at risk or<br>clinically<br>significant<br>impairment |
| Luedke et al. (2024) | USA<br>(CDC) | PASC<br>clinic | 331.11<br>(240.54) | 54 (35/19) | 6/48 | 13.48 (3.1) | Tea-Ch,<br>D-KEFS,<br>WASI-II,<br>WISC-V,<br>WAIS-IV,<br>ChAMP | *n* of sample scoring<br>below 9th<br>percentile,<br>indicating at risk or<br>clinically<br>significant<br>impairment |
| Ng et al.<br>(2022) | USA<br>(CDC) | PASC<br>clinic | 280 (178) | 18 (12/6) | 3/15 | 13.28 (2.70) | ChAMP,<br>CVLT-C,<br>WISC-V,<br>SDMT,<br>WAIS-IV,<br>Tea-Ch,<br>NEPSY-II,<br>D-KEFS | *n* of sample scoring<br>in the borderline/<br>clinical range,<br>indicating at risk or<br>clinically<br>significant<br>impairment |
| Scarselli et<br>al. (2023) | Italy<br>(WHO) | Hospital | 30.42-547.50<br>(243.33)** | 50(22/28) | 35/12 | 8-16 (11.5)<br>** | NEPSY-II,<br>CBCL* | *n* of sample scoring<br>in the borderline/<br>clinical range,<br>indicating at risk or<br>clinically<br>significant<br>impairment |
| Frolli et al.<br>(2021) | Italy<br>(WHO) | Secondary<br>School | 91 (N/A) | *PASC group*<br>50 (20/30)<br>*Comparators*<br>50 (19/31) | 25/75 | *Range*<br>12-13 | WISC-IV,<br>BVN-12-18 | *M* and *SD* of<br>participant scores |
| Jason et al.<br>(2023) | USA<br>(CDC) | Social<br>Media<br>recruitment | 168(76.72) | *PASC Group*<br>19 (12/7)<br>*Comparators* | N/A | *PASC /*<br>*Comparator* | DPSQ* | *M* and *SD* of<br>participant scores |
|  |  |  |  | 19 (12/7) |  | 9.98(4.15) /<br>12.74(2.38) |  |  |

*Note.* \* indicates parent-report measure; \*\* indicates studies that reported follow up time or participant age as minimum – maximum (median); \*\*\* indicates studies that reported follow-up time as median (inter quartile range); F = female; M = male; CFS = Chalder Fatigue Scale; Tea-Ch = Test of Everyday Attention for Children; D-KEFS = Delis-Kaplan Executive Function System Test; WASI-II = Wechsler Abbreviated Scale of Intelligence - 2nd Edition; WISC-V = Wechsler Intelligence Scale for Children - 5th Edition; WAIS-IV = Wechsler Adult Intelligence Scale - 4th Edition; ChAMP = Child and Adolescent Memory Profile; CVLT-C = California Verbal Learning Test – Children's Version; SDMT = Symbol Digit Modalities Test; NEPSY-II = Neuropsychological Assessment - Second Edition; CBCL = Child Behaviour Checklist; WISC-IV = Wechsler Intelligence Scale for Children - Fourth Edition; BVN-12-18 = Batteria per la Valutazione Neuropsicologia per L'adolescenza (translation: Neuropsychological Evaluation Battery for Adolescents); DPSQ = DePaul Pediatric Symptom Questionnaire

### Quality Assessment Outcomes

Table 5 provides a summary of outcomes of the quality assessment following the JBI *Critical Appraisal Tool’s* Checklist for Analytical Cross-Sectional Studies (Moola et al., 2017). Item three of the criteria (valid/reliable exposure measure) was deemed not applicable to the extracted studies because the exposure was a condition. COVID-19 and adherence to diagnostic criteria was addressed by item four. Overall, no studies meeting full text screening criteria were found to have high risk of bias.

**Table 5.**
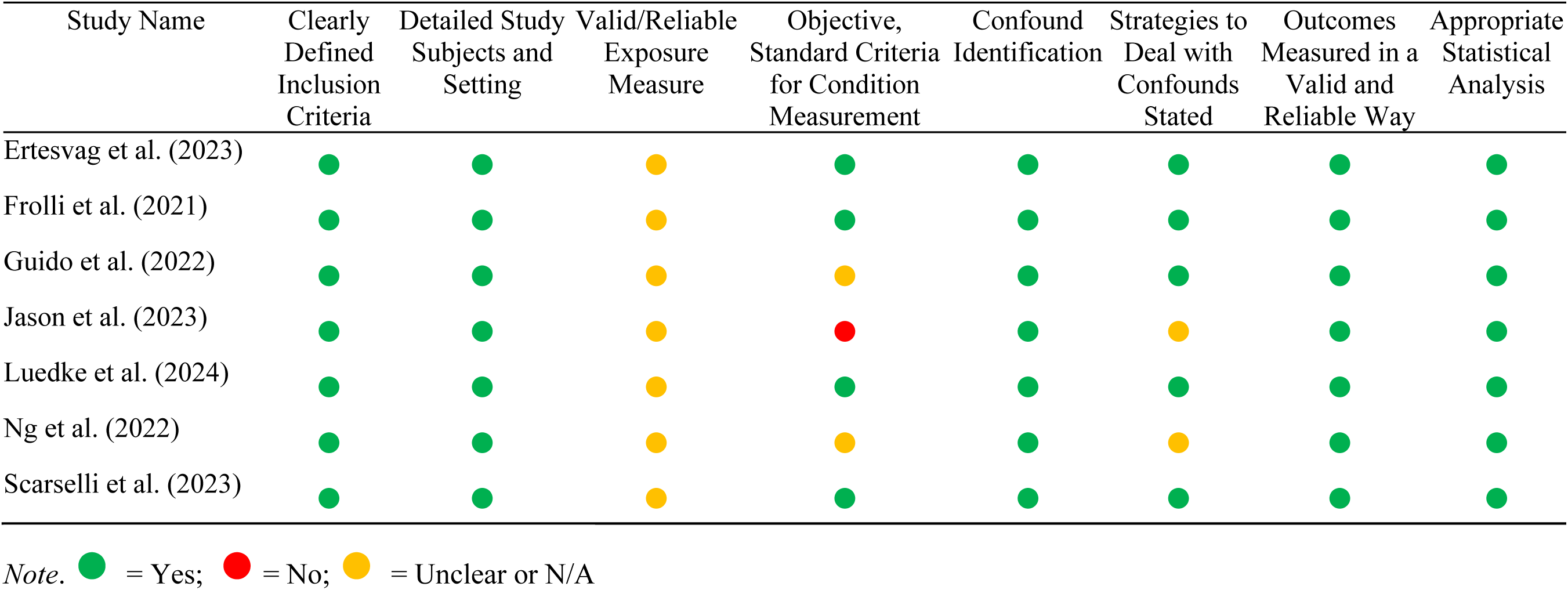
Joanna Briggs Institute Critical Appraisal Tool’s Checklist for Analytical Cross-Sectional Studies.

### Data Coding

All data points (individual items on each outcome measure) were classified in accordance with a specific DSM-5 cognitive domain (American Psychiatric Association, 2013), as shown in Table 6. A clinical neuropsychologist was consulted regarding coding protocols to ensure that classification of each outcome measure reflected current clinical consideration of the cognitive domain being assessed as accurately as possible. A summary of the psychometric tests and report measures used within these studies is provided in Table 7.

**Table 6.**
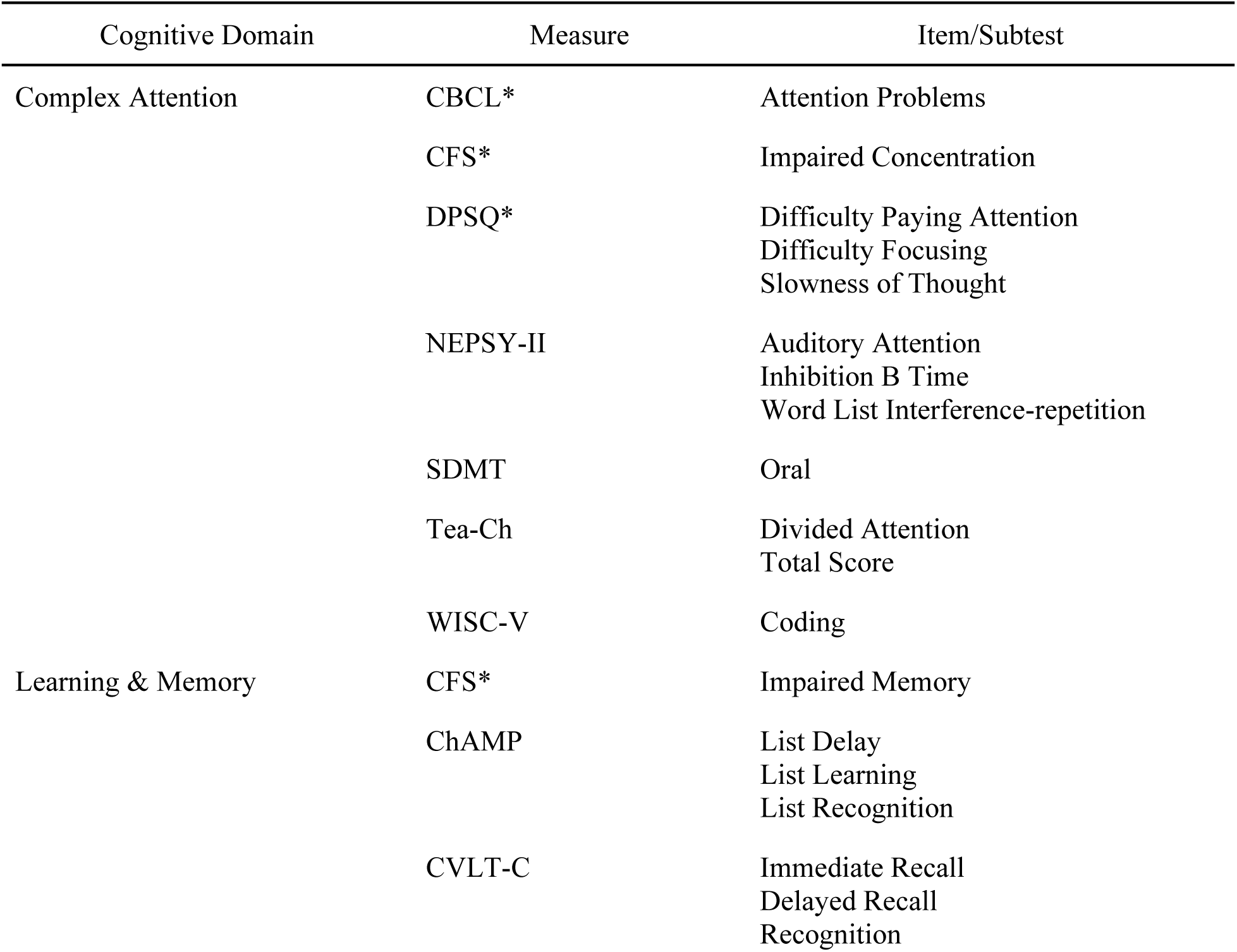
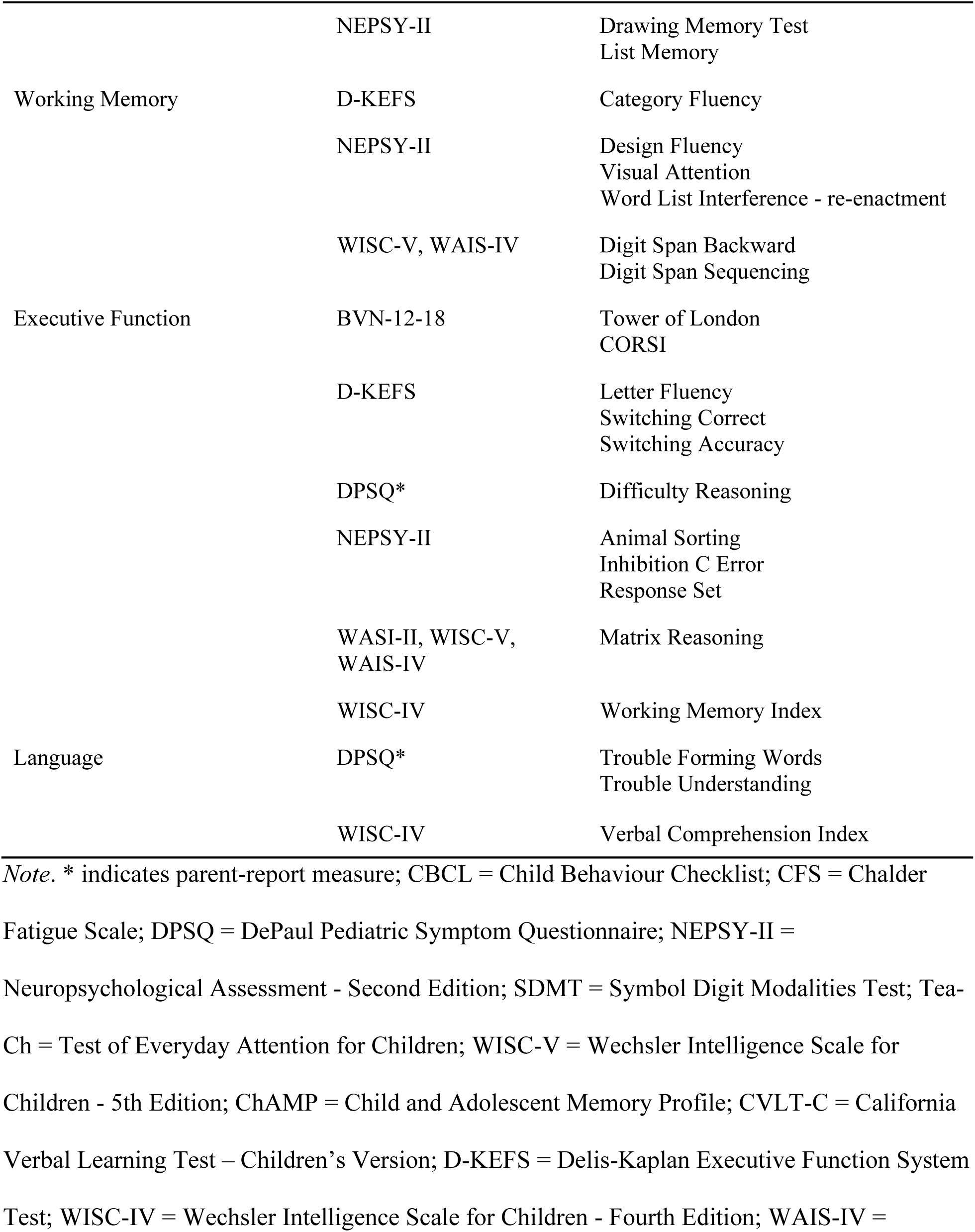
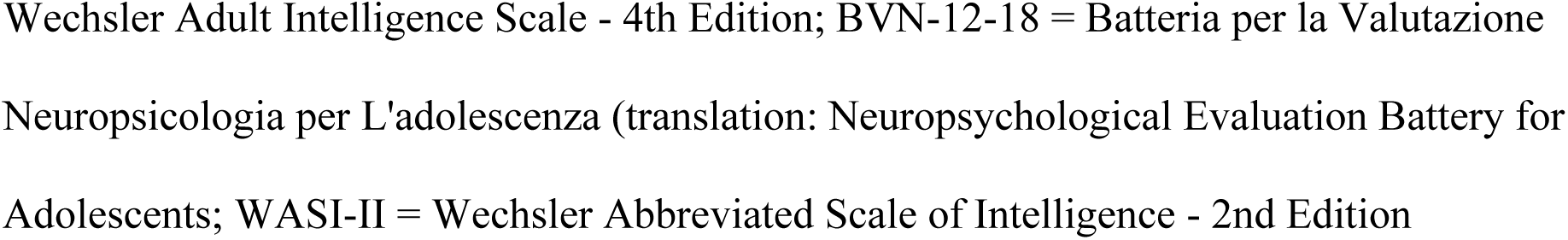
Coding of Each Subtest/Item from Included Studies to DSM-5 Cognitive Domains.

**Table 7.**
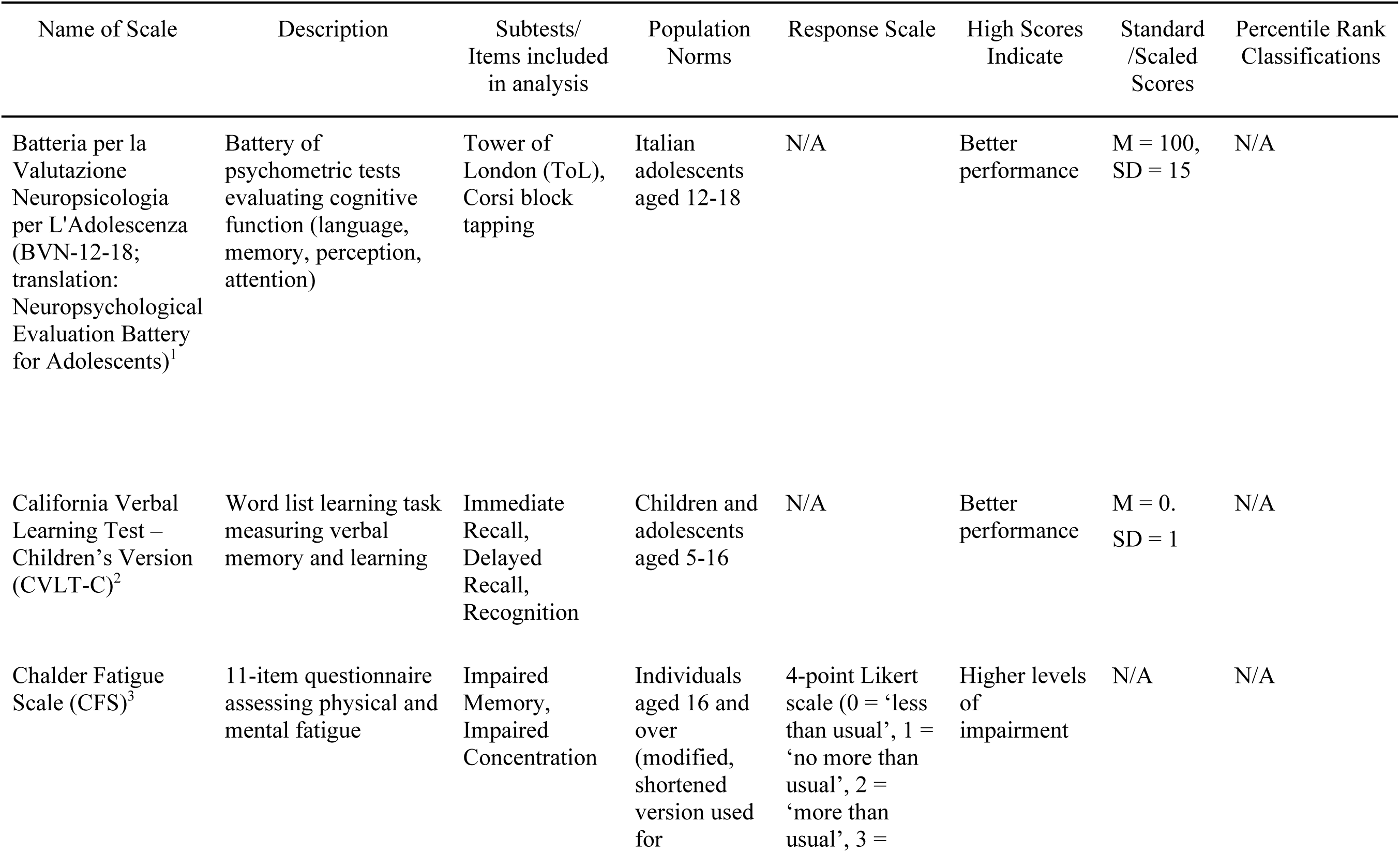
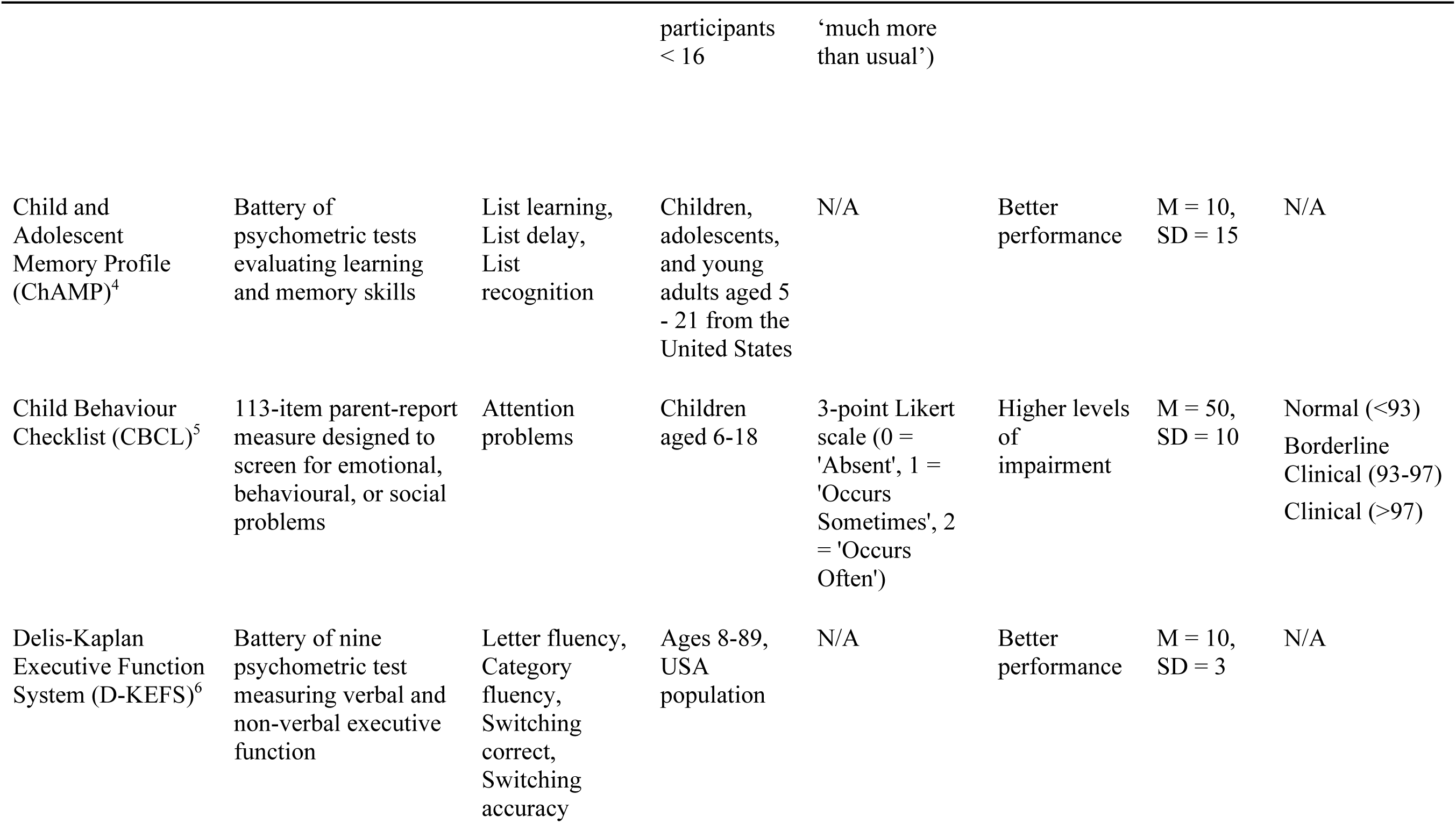
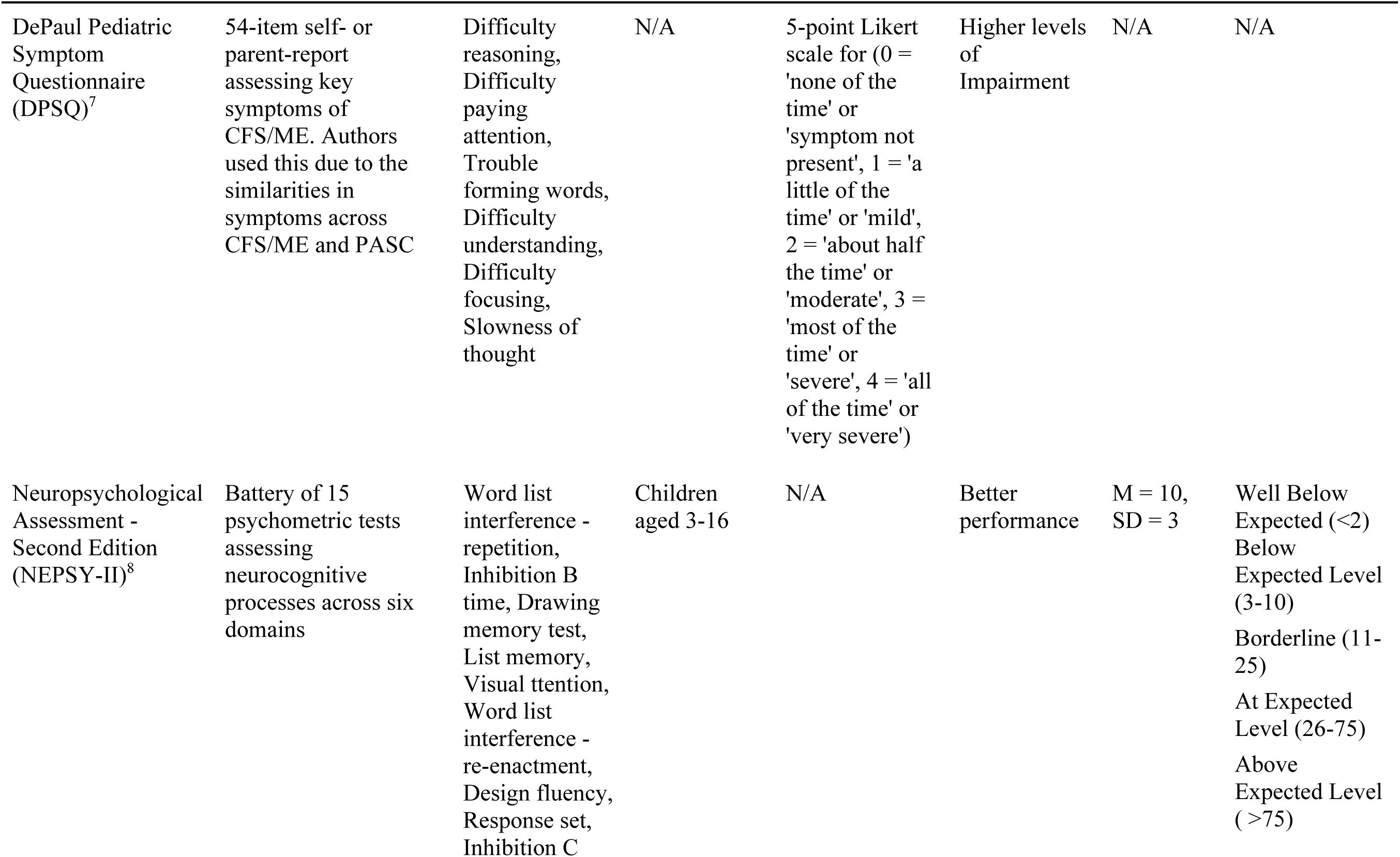
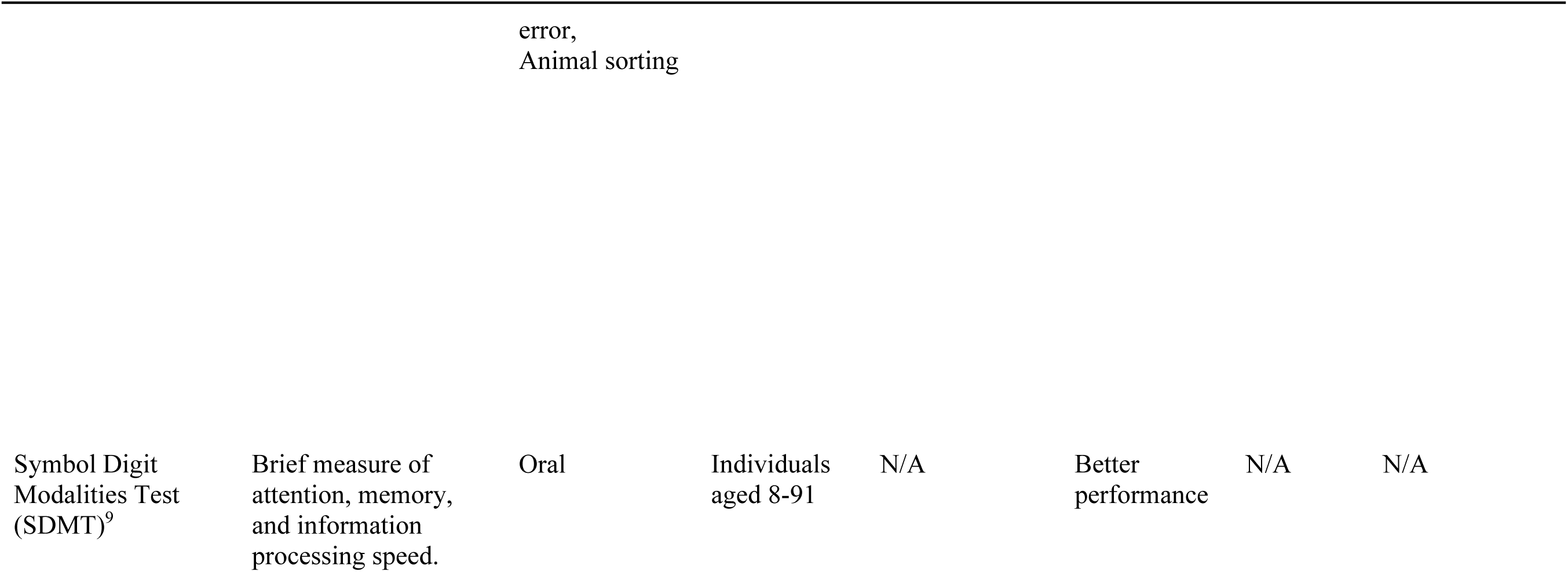
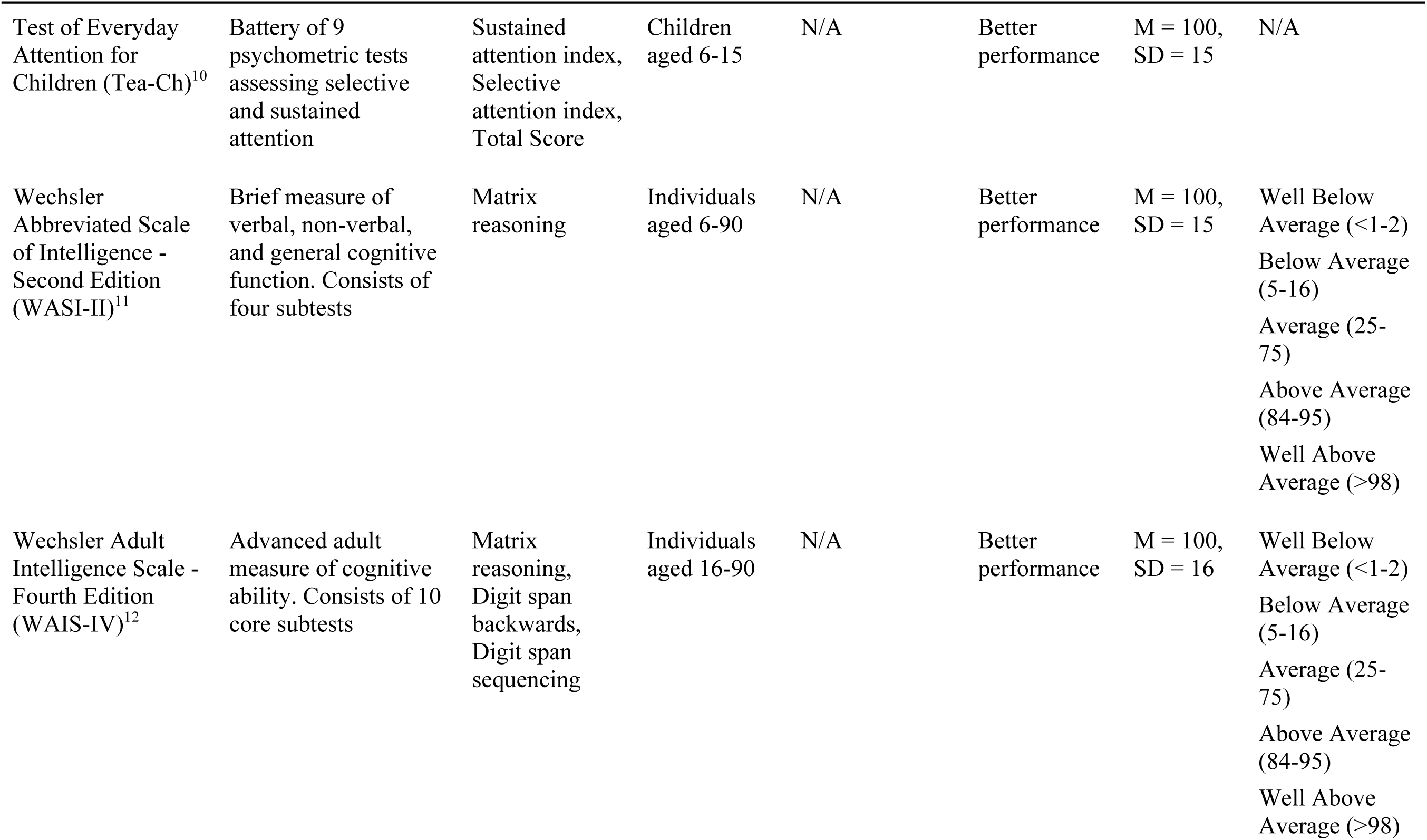
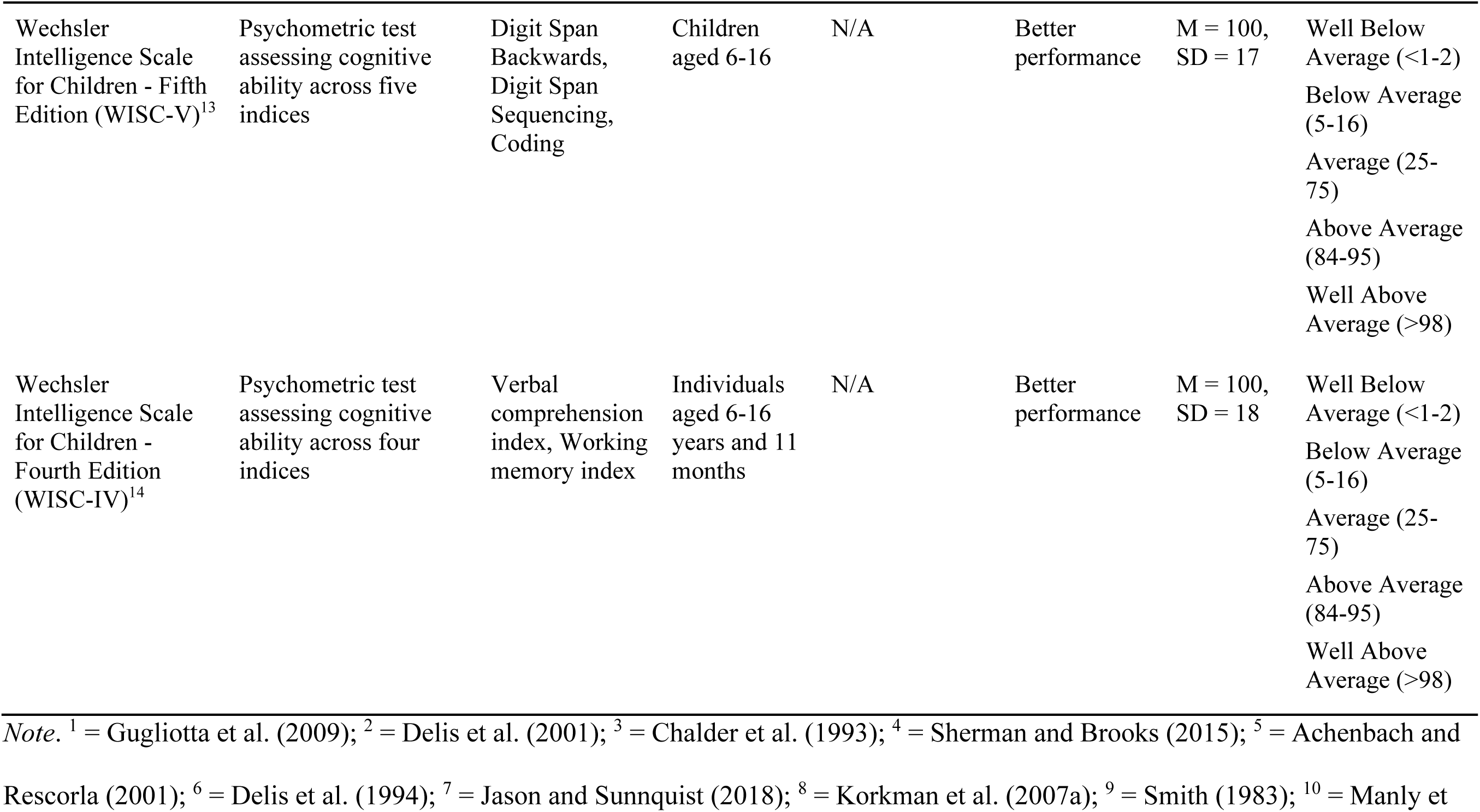
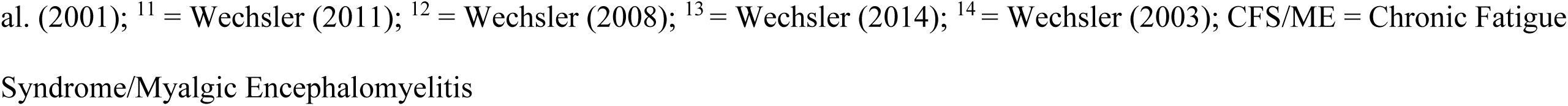
Summary of Psychometric Tests and Reporting Scales used in Selected Studies.

### Meta-Analysis of Prevalence

Of the seven studies selected for inclusion in the analysis, five studies provided 36 data points across 453 participants that were able to be included in the meta-analysis. Pooled prevalence of at risk or clinically significant cognitive impairment was viable for meta-analysis across four of the six DSM-5 cognitive domains. At risk or clinically significant cognitive impairment was determined based on the clinical cut-offs across measures and how these were defined across each study. Despite extracting for hospitalisation status as a potential confounding factor, there was insufficient data available to analyse this effect. Insufficient data were available to run subgroup analyses on group difference-based outcome measurements. Pooled prevalences of at risk or clinically significant impairments observed across complex attention, learning & memory, working memory & executive function are respectively presented below.

#### Complex Attention

Figure 2 shows that 55% of pooled paediatric PASC participants were considered at risk of, or showing clinically significant impairment in complex attention based on parent report or clinical assessment outcomes (95% CI = 39.00% to 71.00%). Heterogeneity was considerable, τ^2^ = .05, SE = .03, *p* < .001, *I*^2^ = 90.90%. The rank correlation test for funnel plot asymmetry indicated no publication bias, Kendall’s τ = .18, *p* = .51. The studies highlighted by asterisks show parents report approximately 25-84% prevalence of at risk or clinically significant complex attention impairment.

**Figure 2.**
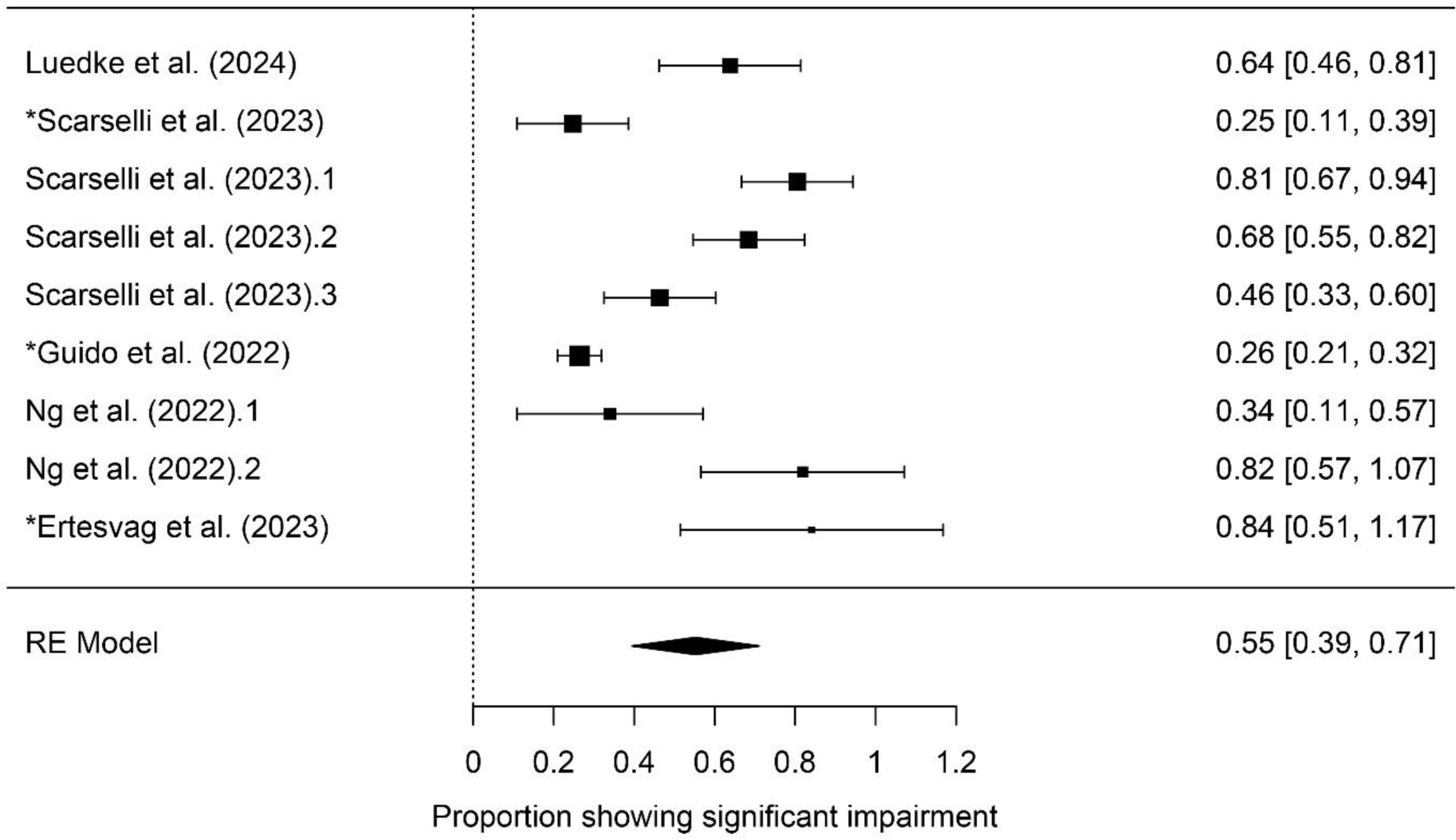
Pooled Prevalence of At Risk or Clinically Significant Complex Attention Impairments in Paediatric PASC Patients. *Note*. * indicates parent-report measure

#### Learning & Memory

Forty-two percent of pooled paediatric PASC participants were considered at risk of, or showing, clinically significant levels of impairment in learning and memory based on clinical assessment outcomes (95% CI = 26.00% to 58.00%) (Figure 3). Heterogeneity was considerable, τ^2^ = .05, SE = .03, p < .001, I^2^ = 86.1%. The rank correlation test for funnel plot asymmetry indicated no publication bias, Kendall’s τ = -.09, p = .74. The studies highlighted by asterisks show parents report approximately 73% prevalence of at risk or clinically significant complex attention impairment.

**Figure 3.**
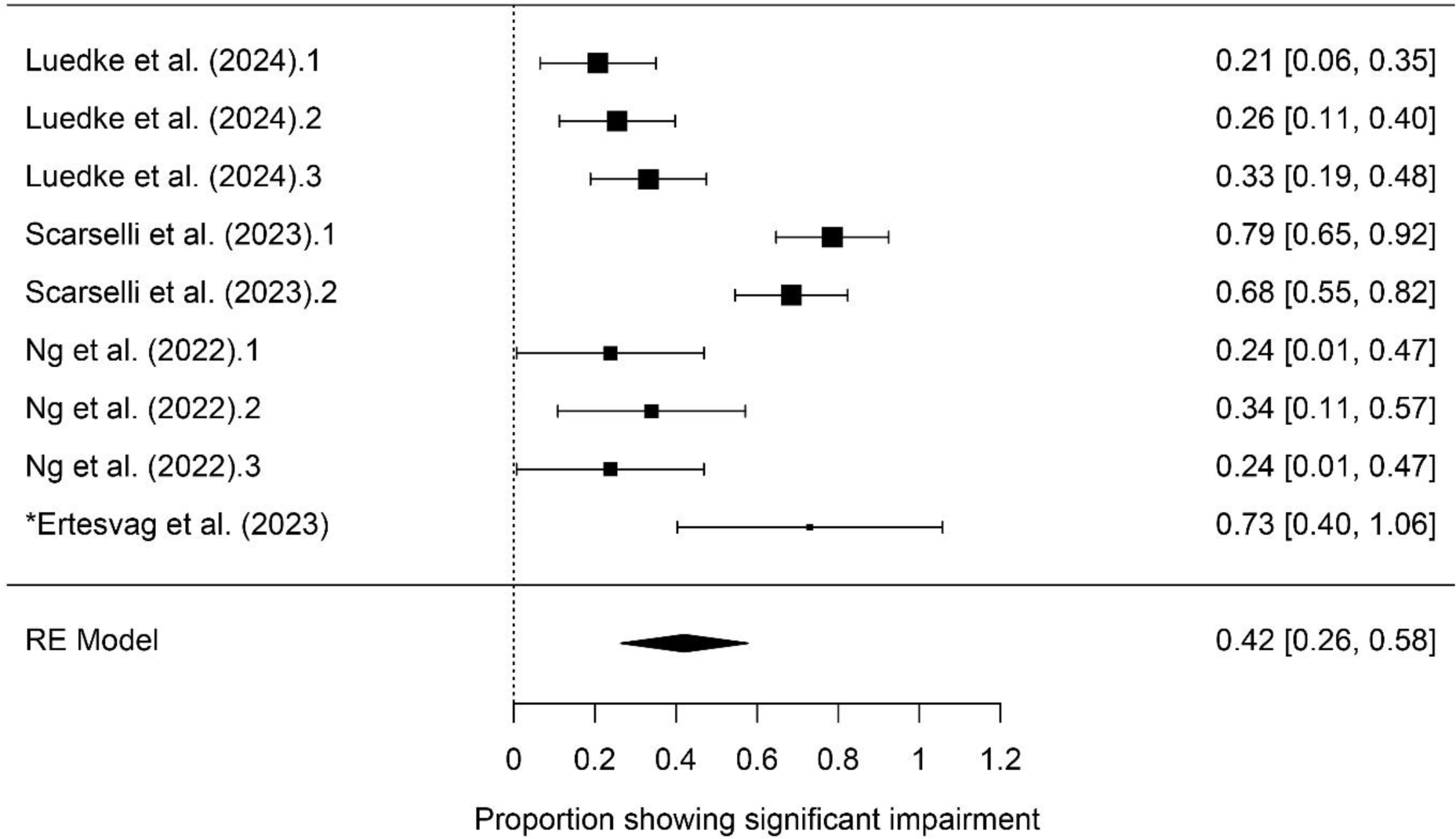
Pooled Prevalence of At Risk or Clinically Significant Learning & Memory Impairments in Paediatric PASC Patients. *Note. * indicates parent report measure*

#### Working Memory

Forty two percent of pooled paediatric PASC participants displayed impairments in working memory that were considered at risk or clinically significant based on clinical assessment outcomes (95% CI = 24.00% to 59.00%) (Figure 4). Heterogeneity was considerable, τ^2^ = .05, SE = .03, *p* < .001, *I*^2^ = 90.31%. The rank correlation test for funnel plot asymmetry indicated no publication bias, Kendall’s τ = .05, *p* = .87.

**Figure 4.**
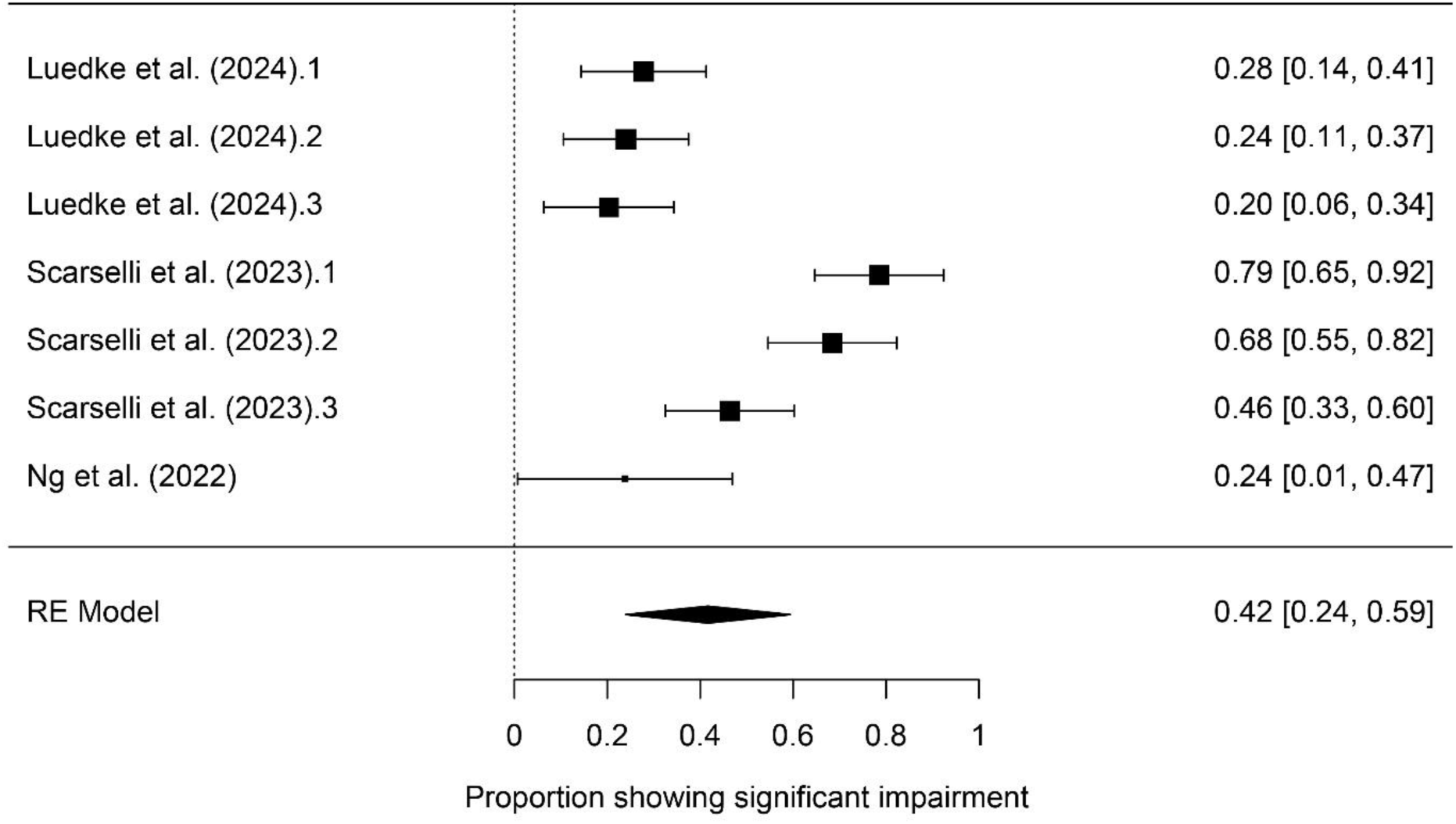
Pooled Prevalence of At Risk or Clinically Significant Working Memory Impairments in Paediatric PASC Patients.

#### Executive Function

Finally, 35% of pooled paediatric PASC participants displayed impairments in executive function that were considered at risk or clinically significant based on clinical assessment outcomes (95% CI = 18.00% to 52.00%) (Figure 5). Heterogeneity was considerable, τ^2^ = .07, SE = .04, *p* < .001, *I*^2^ = 91.47%. The rank correlation test for funnel plot asymmetry indicated no publication bias, Kendall’s τ = -.48, *p* = .07).

**Figure 5.**
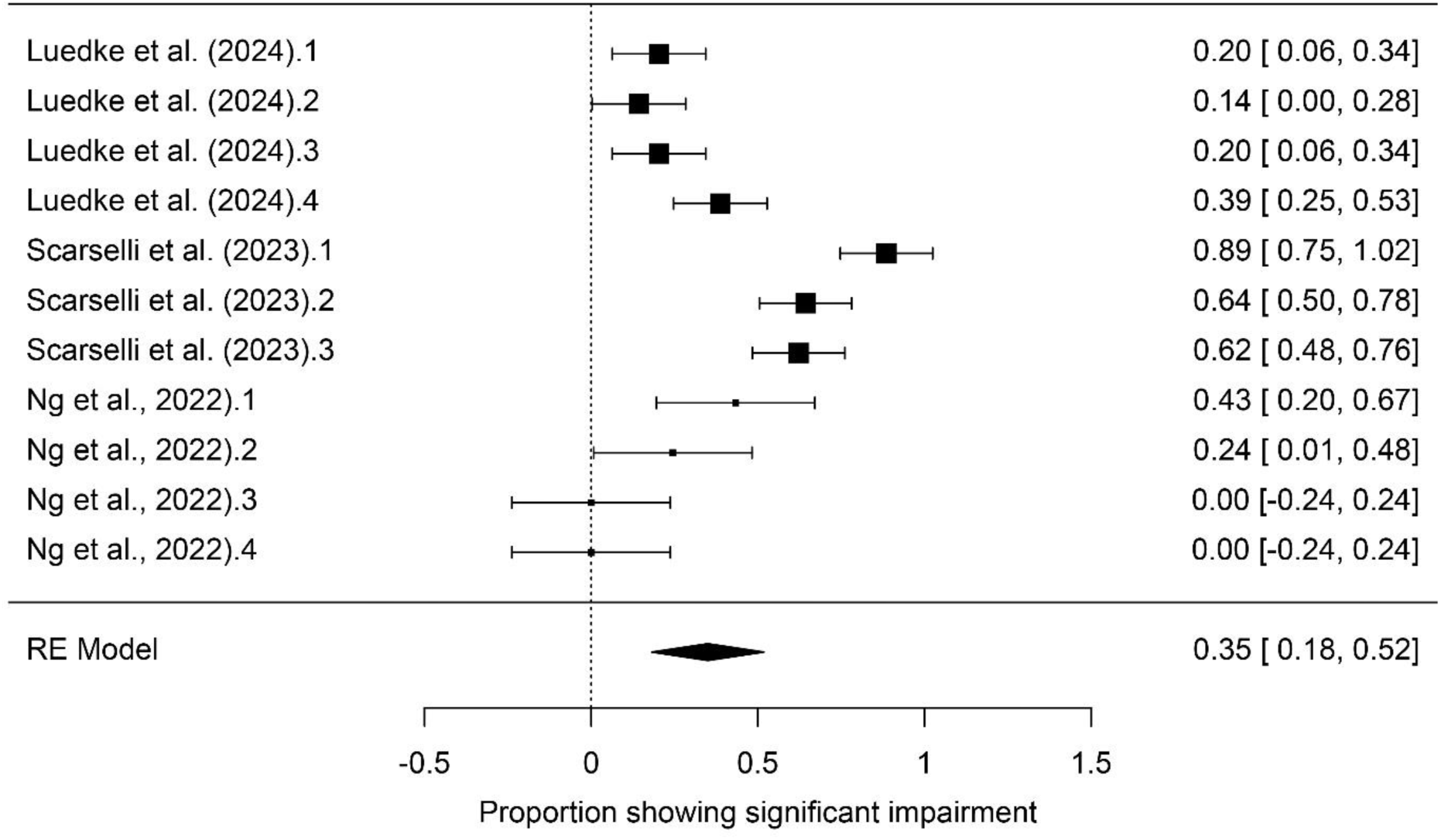
Pooled Prevalence of At Risk or Clinically Significant Executive Function Impairments in Paediatric PASC Patients.

### Narrative Synthesis

Two of the seven included studies could not be included in the prevalence analysis due to the lack of prevalence data reported. Furthermore, these studies did not include comparable control groups, and thus were not viable for meta-analysis of group differences. Frolli et al. (2021) utilised healthy controls while Jason et al. (2023) compared children and adolescents diagnosed with chronic fatigue syndrome/myalgic encephalomyelitis (CFS/ME) to those with PASC. Both studies provided data on complex attention and executive function, and on cognitive impairment in the language domain that were could be qualitatively synthesised.

Both studies provided data on executive functioning and language across the WISC-IV, BVN-12-18 and DPSQ. As visualised in Figure 6, all PASC groups from Frolli et al’s (2021) study were assessed to have impairments in executive function and language, irrespective of hospitalisation status. Hospitalised paediatric PASC patients experienced the highest levels of impairment, supported by large effect sizes across subtests measuring executive function and language as displayed in Table 8.

**Figure 6.**
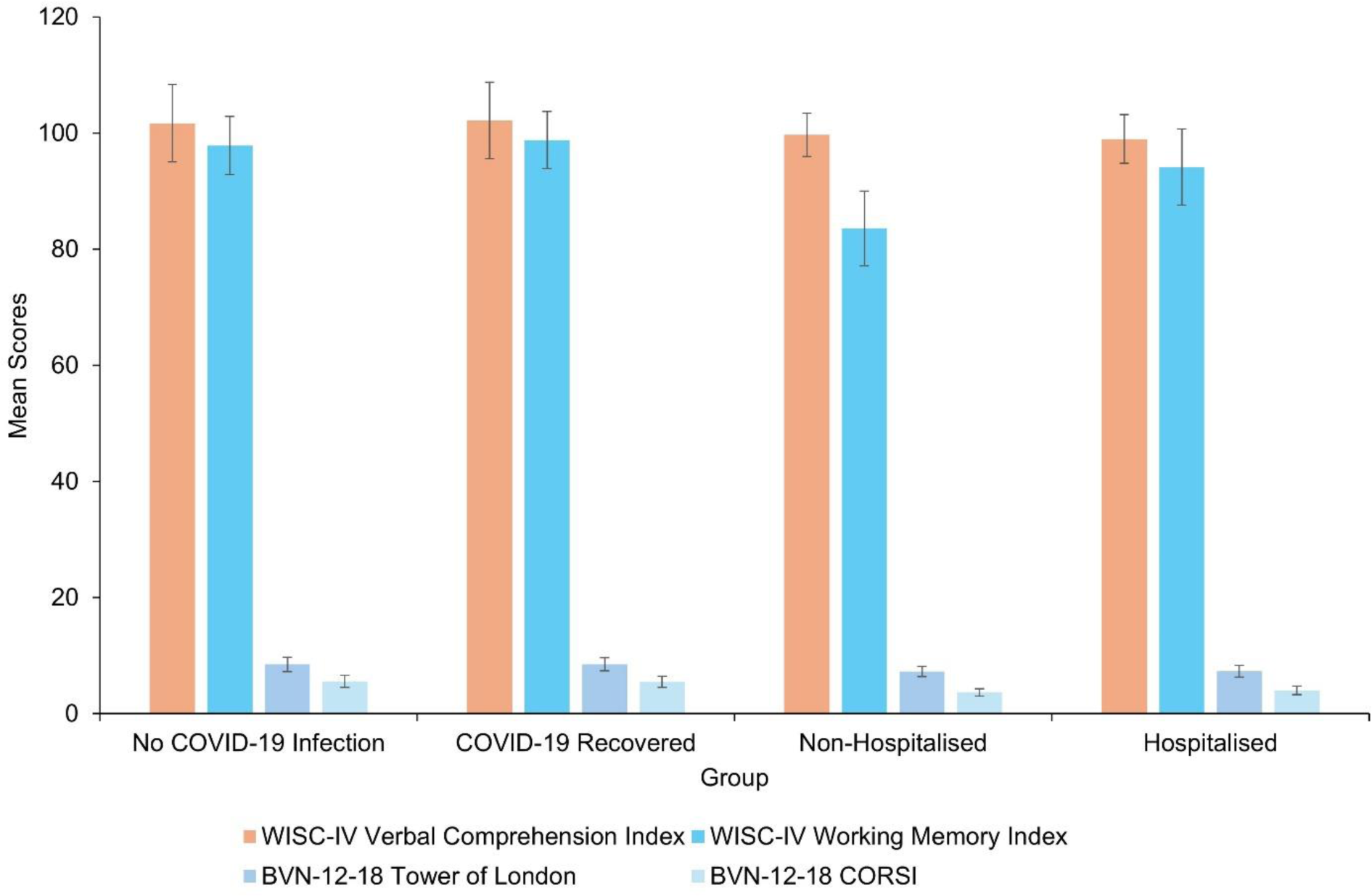
Means and Standard Deviations of Clinical Assessment Scores Across Controls with No Previous COVID-19 Infection, COVID-19 Recovered Participants (with no PASC diagnosis), Non-Hospitalised PASC Patients and Hospitalised PASC Patients (Frolli et al., 2021). *Note*. Orange bars indicate language subtests; blue bars indicate executive function subtests; WISC-IV = Wechsler Intelligence Scale for Children - Fourth Edition; BVN-12-18 = Batteria per la Valutazione Neuropsicologia per L’Adolescenza (translation: Neuropsychological Evaluation Battery for Adolescents)

**Table 8.** Hedge’s g Effect Sizes of Clinical Assessment Scores Between COVID-19 Recovered, Non-Hospitalised PASC and Hospitalised PASC Groups and Controls (Frolli et al., 2021).

| Outcome Measure by Cognitive Domain | COVID-19 Recovered Hedge's g | Non-Hospitalised Hedge's g | Hospitalised Hedge's g |
| --- | --- | --- | --- |
| Executive Function |  |  |  |
| WISC- IV WMI | 0.20 | -0.63 | -2.44 |
| BVN-12-18 |  |  |  |
| Tower of London | 0.00 | -1.02 | -1.10 |
| CORSI | -0.08 | -1.66 | -2.15 |
| Language |  |  |  |
| WISC-IV VCI | 0.07 | -0.48 | -0.37 |

*Note.* WISC-IV = Wechsler Intelligence Scale for Children – Fourth Edition; WMI = Working Memory Index; BVN-12-18 = Batteria per la Valutazione Neuropsicologia per L'Adolescenza (translation: Neuropsychological Evaluation Battery for Adolescents); VCI = Verbal Comprehension Index

Consistent with the findings in the complex attention domain, PASC patients from Jason et al. (2023) exhibit impairments in executive functioning, but not to same extent as CFS/ME patients (*g* = −0.96). Only Jason et al. (2023) provided data points that measured complex attention. As shown in Figure 7, PASC patients displayed difficulties across complex attention items as reported by parents on the DPSQ, but not to the degree that was exhibited by the CFS/ME group. As summarised in Table 9, medium and large effect sizes were observed across complex attention items, notably in difficulty paying attention (*g* = −0.99) and slowness of thought (*g* = −0.51).

**Figure 7.**
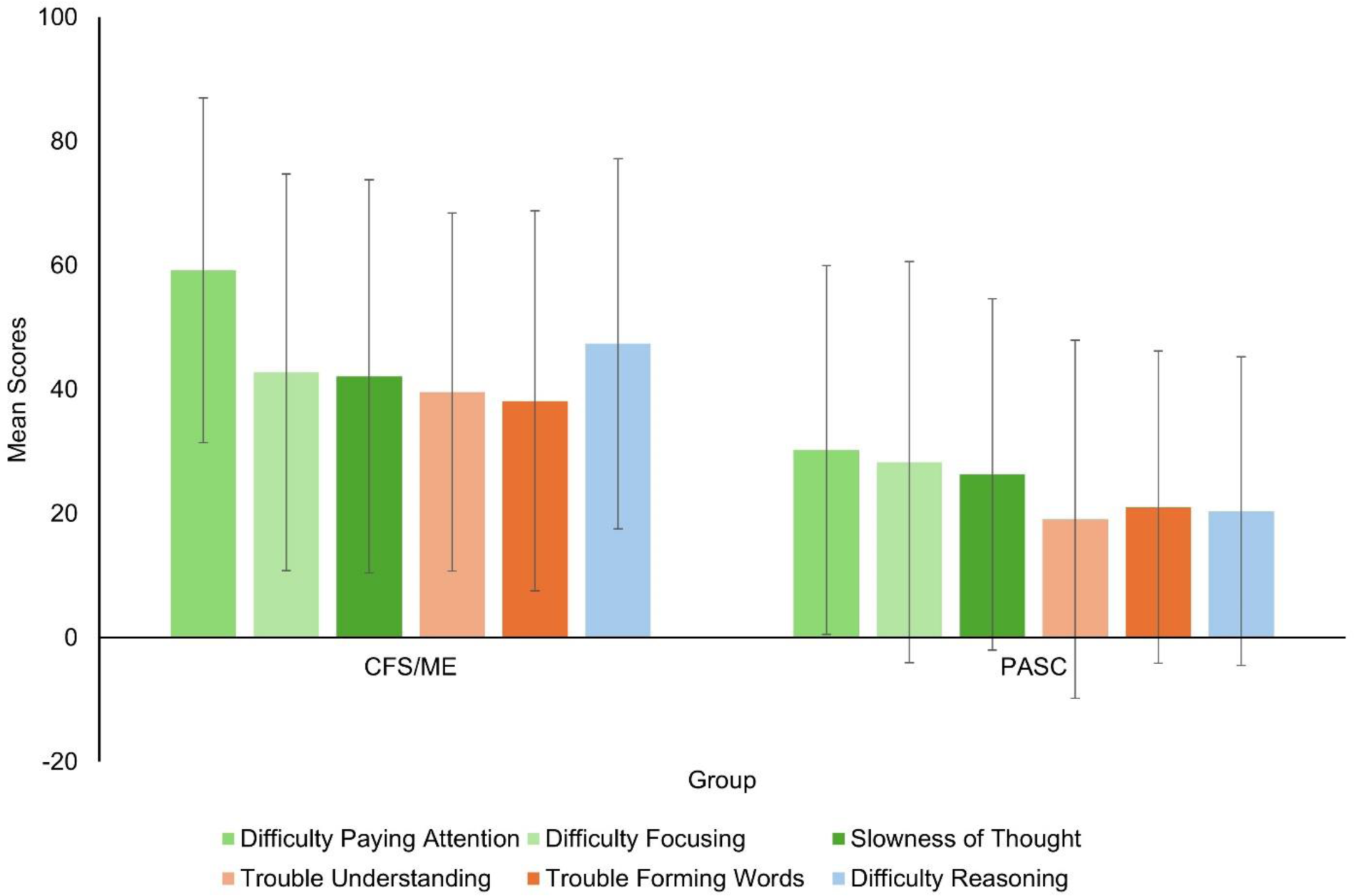
Means and Standard Deviations of Paediatric PASC Participants’ DPSQ Scores compared to Paediatric CFS/ME Patients (Jason et al., 2023). *Note*. Higher mean scores indicate greater difficulties on each item; Blue bars indicate items coded as executive function; orange bars indicate items coded as language; green bars indicate items coded as complex attention; DPSQ = DePaul Symptom Questionnaire.

**Table 9.** Hedges’ g Effect Sizes of DPSQ Scores Between Paediatric Chronic Fatigue Syndrome Group and Paediatric PASC Group (Jason et al., 2023).

| Item on DPSQ by Cognitive Domain | Hedge's g |
| --- | --- |
| Complex Attention |  |
| Difficulty Paying Attention | -0.99 |
| Difficulty Focusing | -0.44 |
| Slowness of Thought | -0.51 |
| Executive Function |  |
| Difficulty Reasoning | -0.96 |
| Language |  |
| Trouble Forming Words | -0.60 |
| Trouble Understanding | -0.78 |

*Note.* DPSQ = DePaul Symptom Questionnaire

## Discussion

The current systematic review and meta-analysis aimed to synthesise literature reporting cognitive impairment associated with PCNS in paediatric PASC populations, and to provide a more nuanced cognitive profile of these impairments based on the DSM-5 cognitive domains such as complex attention, learning and memory, working memory, and executive function. Of the 4271 records screened, seven studies met the inclusion criteria and provided evidence that high proportions of paediatric PASC patients are likely to experience clinically significant deficits across complex attention, learning and memory, working memory, and executive function.

### Prevalence Analysis Outcomes

#### Risk for Impairments in Complex Attention in Paediatric PCNS

Complex attention was found to be the most prevalent cognitive impairment observed across all analysed domains. As such, children with PCNS are likely to present with difficulties in sustaining, holding or dividing attention, impacting the ability to encode information into memory stores, learn new information, and carry out higher order thinking processes (Oberauer, 2019; Rabiner et al., 2016). The findings of the present analysis align with previous studies measuring attention impairment based on psychometric tests, parent-reports and self-reports (Brackel et al., 2021; Buonsenso et al., 2021; Buonsenso et al., 2022; Sterky et al., 2021). This is also in line with recent reports in adult PCNS populations where deficits in several attention indices were observed (Cerioli et al., 2024; Charles James et al., 2025; Delgado-Alonso et al., 2025; Su et al., 2024). Although data were not sufficient to conduct separate analyses for psychometric assessment and parent-, or self-report outcomes, the current findings suggest that the impact of impairments in complex attention are perceived by patients and caregivers, and that reports from these groups are good indicators for the presence of cognitive impairment, though potentially with a lower degree of sensitivity than formal psychometric assessments. This also suggests that development of age-appropriate scales for evaluating PCNS-related impairments in children are warranted to fully characterise the nature of cognitive deficits in this population and provide clinicians with standardised, evidence-based tools to guide more nuanced neuropsychological assessment, and the formulation of appropriate intervention strategies.

#### Risk for Impairments in Learning & Memory in Paediatric PCNS

The finding of a high prevalence of at risk or clinically significant learning and memory impairments is also consistent with previous studies reporting impairments across these subdomains in children (Brackel et al., 2021; Gonzalez-Aumatell et al., 2022) and adults (Charles James et al., 2025; Gonzalez Aleman et al., 2025; Su et al., 2024). Learning and memory impairments often present in children through an inability to recall new information, difficulty following new instructions and often forgetting requirements such as homework (Wiguna et al., 2012) and would not be unexpected if complex attention is compromised, as detailed above. These findings are consistent with the levels of impairment in learning and memory observed in adult PASC populations (Fanshawe et al., 2024). When considering the source of heterogeneity in this analysis, it is interesting to note that a higher percentage of participants from Scarselli et al. (2023) were assessed as having at risk or clinically significant learning and memory impairments compared to participants from Luedke et al. (2024). This may be due to the fact that a higher proportion of Scarselli et al.’s (2023) sample were hospitalised (70%) compared to those from Luedke et al. (2024) (11.11%) and suggests that the degree of paediatric PASC-related impairment may be associated with symptom severity in a similar manner observed in adult populations (Alnefeesi et al., 2020; Ceban et al., 2022; Miskowiak et al., 2023).

#### Risk for Impairments in Working Memory and Executive Function in Paediatric PCNS

Prevalence analysis results were consistent with current literature suggesting working memory impairment and executive functioning deficits across paediatric PASC patients (Gonzalez-Aumatell et al., 2022; Pazukhina et al., 2022; Roge et al., 2021), revealing that 44% of patients were deemed at risk or showing clinically significant degrees of impairment for both domains. Working memory deficits may present in children through difficulties in verbal skills and language comprehension as well as solving mathematical equations (Diamond, 2013; Pickering et al., 2023). Executive functioning impairments in children may present as difficulties in problem solving tasks and complex reasoning, as well as difficulty controlling emotions, behaviours, and thoughts (Diamond, 2013). It is acknowledged that this analysis displayed a high risk of publication bias, suggesting that the prevalence of at risk or clinically significant impairment may be inflated. However, our finding of executive functioning impairments in paediatric patients are also in line with those observed in adult populations, suggesting consistency in PCNS symptomology across age groups (Fanshawe et al., 2024).

### Narrative Synthesis Outcomes

Narrative synthesis supported the prevalence analysis findings, suggesting a high incidence of cognitive impairments across both hospitalised and non-hospitalised PASC patients compared to controls who had never contracted COVID-19 or had recovered from infection (Frolli et al., 2021). Effect sizes of hospitalised PASC patients versus control for executive functioning measures were larger than those for non-hospitalised patients, suggesting that COVID-19 symptom severity may have an impact on the severity of cognitive symptoms.

Despite insufficient data to perform any analysis on the language domain, there were medium effect sizes observed in the VCI scores in both non-hospitalised and hospitalised PASC patients, suggesting levels of impairment in this domain. The observation of greater impairments for previously hospitalised patients is in contrast to meta-analyses by Ceban et al., (2022) and Chen et al., (2022) who found no differences in the level of cognitive impairment in hospitalised versus non-hospitalised adults. However, both of investigations included data from samples ranging from 90 days to beyond one year at follow-up, and broader quality of life and cognitive screening tools than those included in the current meta-analysis. The evidence from our narrative synthesis is consistent with outcomes from a recent systematic narrative review reporting case study evidence for an association between viral load, physical symptom severity and cognitive impairments in paediatric patients (Avittan & Kustovs, 2023).

Narrative synthesis also found that paediatric PASC patients show complex attention difficulties, though not to the same extent as a CFS/ME comparator group. This is of interest as CFS/ME and PASC have extremely similar inflammatory presentations (Jason et al., 2023), and the DPSQ is a measure designed to characterise the frequency and severity of CFS/ME symptoms (Jason & Sunnquist, 2018). Given this similarity in presentation, these results demonstrate that the DPSQ may be a useful screening tool for characterising PASC-related cognitive impairment and tracking the chronicity of such impairments over time. Relating DPSQ scores for PASC patients to CFS/ME profiles may provide an indication of the longer-term cognitive impact and ongoing symptom escalation. When considered in conjunction with reports that approximately 44% of hospitalised paediatric patients show neurological involvement (for example, migraine, seizures or stroke) and 25% are diagnosed with Multisystem Inflammatory Syndrome (Kwan et al., 2024), and evidence that prior vaccination status may not predict PCNS impairment (Mukherjee et al., 2025), this again highlights the need for clear patient stratification, appropriate screening tools and a systems-based perspective in understanding the interplay between the broad physiological impact and the neurological/neuropsychological phenotype in PCNS.

### Theoretical Implications

Despite being unable to meta-analyse impairments in language, perceptual-motor function and social cognition domains, the current study has demonstrated that paediatric PASC patients are a risk of impairment in foundational cognitive functions. It is also evident that the magnitude of impairment may be associated with the severity of the infection and subsequent impact of an ongoing elevated immune and physiological response (Wijeratne & Crewther, 2020). By pinpointing complex attention, learning and memory, working memory and executive function as the most prevalent domains impacted, these findings can help inform future symptom management and intervention strategies at a clinical level.

At an educational level, the findings from this study can further inform teachers on what cognitive impairments a child or adolescent with PASC may present with, and subsequently inform better supports in the classroom. This can lead to a more holistic approach to treating and managing paediatric PASC that acknowledges cognitive impairment alongside other symptoms like chronic fatigue, brain fog, and respiratory symptoms (Chen et al., 2023; Wijeratne & Crewther, 2020; World Health Organisation, 2023).

### Limitations

#### Limitations in Current Paediatric PCNS Research

There are many limitations observed across the wider body of PASC literature that precluded more in-depth meta-analysis of the paediatric PCNS profile. Despite collecting data on hospitalisation status and sex differences in participant characteristics, many studies fail to account these variables in their analysis in order to provide a more subtle profile of cognitive impairment. Outcome measures used across included studies varied greatly, preventing further subgroup analyses to identify differences in what is reported by parents or patients and what is detected by clinical assessment. The lack of case-controlled studies is another limitation of the current state of literature available to meta-analyse. Case controlled studies provide a baseline to observe cognitive impairments in a neurotypical population with no COVID-19 diagnosis and can provide points of comparison, increasing the reliability of results (Lewallen & Courtright, 1998).

#### Limitations of the Current Study

In light of the issues described above, the current systematic review and meta-analysis is limited by the small number of studies and extracted data points and as a result, the current analysis was unable to provide prevalence results for language, social cognition and perceptual-motor function domains. Given the hierarchical structure of cognition, it can be assumed that these domains would be impacted due to the high prevalences of complex attention and memory deficits (Fisher, 2019; Harvey, 2019). Included studies were conducted in either the United States of America or Italy, limiting the generalisability of the analysis results due to the overrepresentation of Western countries in the sample. There is evidence suggesting that race or ethnicity can have an impact on PASC symptoms, given the socio-economic and cultural differences influencing healthcare access and vaccination rates (Jassat et al., 2023).

Further, generalisability of these findings may also be threatened by the substantial heterogeneity observed across all prevalence analyses, suggesting that prevalences could be influenced by other risk factors. Variability across participant cohorts was a large limitation of the current study, with insufficient data from included studies available to run subgroup analyses to stratify participant characteristics such as sex, socio-economic status, hospitalisation, COVID-19 vaccination history and symptom severity. These are all moderators that have been seen to influence PASC symptomology, specifically cognitive change, in adults (Jassat et al., 2023; Miller et al., 2024; Watanabe et al., 2023).

### Future Directions

Despite the acknowledged limitations, there are several avenues available for future research. As evidenced by the lack of data available to meta-analyse, future research should investigate PCNS-related impairments across language, perceptual motor-function, and social cognition domains to continue to develop a cognitive profile of impairments.

Future research and meta-analyses can benefit from analysing the differences between the types of outcome measures utilised in identifying PCNS-related cognitive symptoms, as well as the source (informant or clinician). Future research should additionally investigate the impacts of risk factors such as hospitalisation status (as observed through narrative synthesis), COVID-19 symptom severity, and sex, all factors that has established impacts on PASC symptomology in adult populations (Jassat et al., 2023; Masserini et al., 2025; Miller et al., 2024; Narayanan et al., 2025; Watanabe et al., 2023; Wulf Hanson et al., 2022; Zeraatkar et al., 2024; Zhang et al., 2025). Further sub-group investigations comparing younger children with adolescents can provide insights into potential differences in symptom presentations, as suggested by Miller et al. (2024).

Due to the lack of longitudinal data available given the recency of PCNS, future research will benefit from exploring the longitudinal impacts of PCNS on symptom progression and resolution. There is little data on the impacts of PASC years after initial infection, given that the longest follow-up from the studies in this analysis was conducted at 18 months post-infection (Scarselli et al., 2023). Focusing future research here is important for understanding probable symptom trajectories and will further inform time-bound supports a child or adolescent may need based on potential short-term or long-term impacts. Given the findings from the narrative synthesis illustrating similarities between PASC and CFS/ME cohorts due to their comparable inflammatory presentations, management strategies shown to mitigate the symptoms of CFS/ME could further be investigated in their efficacy of mitigating symptoms of PCNS.

### Conclusion

In conclusion, the findings from the current systematic review and meta-analysis further details the cognitive symptoms experienced by paediatric PCNS patients. Despite high heterogeneity, results highlight complex attention, learning and memory, working memory, and executive function as the primary domains likely to show clinically significant levels of impairment. By providing a snapshot of the specific cognitive domains impacted, better awareness of the high likelihood of cognitive impairment as a component of PCNS symptomology can be raised. This can subsequently promote early detection of specific cognitive symptoms to aid children and their parents in accessing interventions, leading to improved clinical outcomes.

## Data Availability

All data produced in the present work are contained in the manuscript.

## Acknowledgments

The Authors would like to thank Dr Hayley Pickering for her assistance in reviewing classification of Neuropsychological Assessment tools in line with cognitive domains.

## List of Abbreviations

|  |  |
| --- | --- |
| ACSN | Australian COVID-19 Serosurveillance Network |
| APA | American Psychological Association |
| BVN 12-18 | Batteria per la Valutazione Neuropsicologia per L'Adolescenza<br>(Neuropsychological Assessment Battery for Adolescents) |
| CBCL | Child Behaviour Checklist |
| CDC | Centre for Disease Control |
| CFS/ME | Chronic Fatigue Syndrome/Myalgic Encephalomyelitis |
| ChAMP | Child and Adolescent Memory Profile |
| CI | Confidence Intervals |
| COVID-19 | Coronavirus Disease 2019 |
| D-KEFS | Delis-Kaplan Executive Function System Test |
| DPSQ | DePaul Pediatric Symptom Questionnaire |
| DSM-5 | Diagnostic and Statistical Manual of Mental Disorders - 5th Edition |
| MAJOR | Meta-Analysis for Jamovi |
| MoCA | Montreal Cognitive Assessment |
| NEPSY-II | Neuropsychological Assessment - Second Edition |
| PASC | Post-Acute Sequelae of COVID-19 |
| PCC | Post-Covid Condition |
| PCNS | Post-COVID-19 Neurological Syndrome |
| PRISMA | Preferred Reporting Items for Systematic Reviews and Meta-Analyses |
| PSC | Paediatric Symptom Checklist |
| SARS-CoV-2 | Severe Acute Respiratory Syndrome Coronavirus 2 |
| Tea-Ch | Test of Everyday Attention for Children |
| ToL | Tower of London subtest |
| WAIS-IV | Wechsler Adult Intelligence Scale - 4th Edition |
| WASI-II | Wechsler Abbreviated Scale of Intelligence - 2nd Edition |
| WHO | World Health Organisation |
| WISC-IV | Wechsler Intelligence Scale for Children - 4th Edition |
| WISC-V | Wechsler Intelligence Scale for Children - 5th Edition |

## References

Achenbach, T. M. (1991). Integrative Guide for the 1991 CBCL/4-18, YSR, and TRF Profiles. University of Vermont.

Achenbach, T. M., & Rescorla, L. A. (2001). *Manual for the ASEBA School-age Forms & Profiles: An Integrated System of Multi-informant Assessment*. Research Center for Children, Youth & Families.

Adolphs, R. (2001). The neurobiology of social cognition. Current Opinion in Neuropsychology, 11(2), 231–239. 10.1016/s0959-4388(00)00202-6

Alghamdi, H. Y., Alrashed, A. M., Jawhari, A. M., & Abdel-Moneim, A. S. (2022). Neuropsychiatric symptoms in post-COVID-19 long haulers. Acta Neuropsychiatrica, 34(6), 318–329. 10.1017/neu.2022.13

Alnefeesi, Y., Siegel, A., Lui, L. M. W., Teopiz, K. M., Ho, R. C. M., Lee, Y., Nasri, F., Gill, H., Lin, K., Cao, B., Rosenblat, J. D., & McIntyre, R. S. (2020). Impact of SARS-CoV-2 Infection on Cognitive Function: A Systematic Review. Frontiers in Psychiatry, 11, 621773. 10.3389/fpsyt.2020.621773

Alwan, N. A., & Johnson, L. (2021). Defining long COVID: Going back to the start. Med, 2(5), 501–504. 10.1016/j.medj.2021.03.003

American Psychiatric Association. (2013). Diagnostic and statistical manual of mental disorders: DSM-5 (5th ed.). American Psychiatric Association.

Arain, M., Haque, M., Johal, L., Mathur, P., Nel, W., Rais, A., Sandhu, R., & Sharma, S. (2013). Maturation of the adolescent brain. Neuropsychiatric Disease and Treatment, 9, 449–461. 10.2147/NDT.S39776

Ashkenazi-Hoffnung, L., Shmueli, E., Ehrlich, S., Ziv, A., Bar-On, O., Birk, E., Lowenthal, A., & Prais, D. (2021). Long COVID in Children: Observations From a Designated Pediatric Clinic. Pediatric Infectious Disease Journal, 40(12), e509–e511. 10.1097/INF.0000000000003285

Australian COVID-19 Serosurveillance Network. (2023). Seroprevalence of SARS-CoV-2-specific antibodies among Australian Blood Doners – Round 4. University of New South Wales Kirby Institute. https://www.kirby.unsw.edu.au/sites/default/files/documents/COVID19-Blood-Donor-Report-Round4-Nov-Dec-2022%5B1%5D.pdf

Australian National Health and Medical Research Centre. (2019). Assessing risk of bias. https://www.nhmrc.gov.au/guidelinesforguidelines/develop/assessing-risk-bias

Avittan, H., & Kustovs, D. (2023). Cognition and Mental Health in Pediatric Patients Following COVID-19. International Journal of Environmental Research and Public Health, 20(6). 10.3390/ijerph20065061

Bonilla, H., Peluso, M. J., Rodgers, K., Aberg, J. A., Patterson, T. F., Tamburro, R., Baizer, L., Goldman, J. D., Rouphael, N., Deitchman, A., Fine, J., Fontelo, P., Kim, A. Y., Shaw, G., Stratford, J., Ceger, P., Costantine, M. M., Fisher, L., O’Brien, L.,…McComsey, G. A. (2023). Therapeutic trials for long COVID-19: A call to action from the interventions taskforce of the RECOVER initiative. Front Immunol, 14, 1129459. 10.3389/fimmu.2023.1129459

Brackel, C. L. H., Lap, C. R., Buddingh, E. P., van Houten, M. A., van der Sande, L., Langereis, E. J., Bannier, M., Pijnenburg, M. W. H., Hashimoto, S., & Terheggen-Lagro, S. W. J. (2021). Pediatric long-COVID: An overlooked phenomenon? Pediatric Pulmonology, 56(8), 2495–2502. 10.1002/ppul.25521

Bramer, W. M., Rethlefsen, M. L., Kleijnen, J., & Franco, O. H. (2017). Optimal database combinations for literature searches in systematic reviews: a prospective exploratory study. Systematic Reviews, 6(1), 245. 10.1186/s13643-017-0644-y

Bull-Otterson, L., Baca, S., Saydah, S., Boehmer, T. K., Adjei, S., Gray, S., & Harris, A. M. (2022). Post–COVID Conditions Among Adult COVID-19 Survivors Aged 18–64 and ≥65 Years — United States, March 2020–November 2021. Morbidity and Mortality Weekly Report, 71(21), 713-717. 10.15585/mmwr.mm7121e1

Buonsenso, D., Munblit, D., De Rose, C., Sinatti, D., Ricchiuto, A., Carfi, A., & Valentini, P. (2021). Preliminary evidence on long COVID in children. Acta Paediatrica, 110(7), 2208–2211. 10.1111/apa.15870

Buonsenso, D., Pazukhina, E., Gentili, C., Vetrugno, L., Morello, R., Zona, M., De Matteis, A., D’Ilario, F., Lanni, R., Rongai, T., Del Balzo, P., Fonte, M. T., Valente, M., De Rose, C., Munblit, D., Sigfrid, L., & Valentini, P. (2022). The Prevalence, Characteristics and Risk Factors of Persistent Symptoms in Non-Hospitalized and Hospitalized Children with SARS-CoV-2 Infection Followed-Up for up to 12 Months: A Prospective, Cohort Study in Rome, Italy. Journal of Clinical Medicine, 11(22). 10.3390/jcm11226772

Ceban, F., Ling, S., Lui, L. M. W., Lee, Y., Gill, H., Teopiz, K. M., Rodrigues, N. B., Subramaniapillai, M., Di Vincenzo, J. D., Cao, B., Lin, K., Mansur, R. B., Ho, R. C., Rosenblat, J. D., Miskowiak, K. W., Vinberg, M., Maletic, V., & McIntyre, R. S. (2022). Fatigue and cognitive impairment in Post-COVID-19 Syndrome: A systematic review and meta-analysis. *Brain*, Behaviour and Immunity, 101, 93–135. 10.1016/j.bbi.2021.12.020

Cerioli, M., Giacovelli, L., Nostro, C., Larini, L., Castiglioni, M., Scarpa, C., Cassina, N., Nicotra, A., Maestri, G., Cucumo, V., Masserini, F., Pomati, S., Cirnigliaro, G., Pantoni, L., & Dell’Osso, B. (2024). Post-COVID condition: a focus on psychiatric symptoms and diagnoses in patients with cognitive complaints. CNS Spectr, 1-7. 10.1017/S1092852924000464

Chalder, T., Berelowitz, G., Pawlikowska, T., Watts, L., Wessely, S., Wright, D., & Wallace, E. P. (1993). Development of a fatigue scale. J Psychosom Res, 37(2), 147–153. 10.1016/0022-3999(93)90081-p

Charles James, J., Schulze, H., Siems, N., Prehn, C., Quast, D. R., Trampe, N., Gold, R., & Faissner, S. (2025). Neurological post-COVID syndrome is associated with substantial impairment of verbal short-term and working memory. Sci Rep, 15(1), 1695. 10.1038/s41598-025-85919-x

Chen, C., Haupert, S. R., Zimmermann, L., Shi, X., Fritsche, L. G., & Mukherjee, B. (2022). Global Prevalence of Post-Coronavirus Disease 2019 (COVID-19) Condition or Long COVID: A Meta-Analysis and Systematic Review. Journal of Infectious Diseases, 226(9), 1593–1607. 10.1093/infdis/jiac136

Chen, E. Y., Burton, J. M., Johnston, A., Morrow, A. K., Yonts, A. B., & Malone, L. A. (2023). Considerations in Children and Adolescents Related to Coronavirus Disease 2019 (COVID-19). Physical Medicine and Rehabilitation Clinics of North America, 34(3), 643–655. 10.1016/j.pmr.2023.03.004

Choudhury, N. A., Mukherjee, S., Singer, T., Venkatesh, A., Perez Giraldo, G. S., Jimenez, M., Miller, J., Lopez, M., Hanson, B. A., Bawa, A. P., Batra, A., Liotta, E. M., & Koralnik, I. J. (2025). Neurologic Manifestations of Long COVID Disproportionately Affect Young and Middle-Age Adults. Ann Neurol, 97(2), 369–383. 10.1002/ana.27128

Cohen, J. (1988). Statistical power analysis for the behavioural sciences (2nd ed.). Lawrence Erlbaum Associates.

Daffner, K. R., Gale, S. A., Barrett, A. M., Boeve, B. F., Chatterjee, A., Coslett, H. B., D’Esposito, M., Finney, G. R., Gitelman, D. R., Hart, J. J., Jr., Lerner, A. J., Meador, K. J., Pietras, A. C., Voeller, K. S., & Kaufer, D. I. (2015). Improving clinical cognitive testing. Neurology, 85(10), 910–918. 10.1212/WNL.0000000000001763

Dams-O’Connor, K., & Gordon, W. A. (2013). Integrating interventions after traumatic brain injury: A synergistic approach to neurorehabilitation. Brain Impairment, 14(1), 51–62. 10.1017/BrImp.2013.9

Davis, H. E., Assaf, G. S., McCorkell, L., Wei, H., Low, R. J., Re’em, Y., Redfield, S., Austin, J. P., & Akrami, A. (2021). Characterizing long COVID in an international cohort: 7 months of symptoms and their impact. EClinicalMedicine, 38, 101019–101019. 10.1016/j.eclinm.2021.101019

Deeks, J. J., Higgins, J. P. T., & Altman, D. G. (2023). Analysing data and undertaking meta-analyses. In J. P. T. Higgins, J. Thomas, J. Chandler, M. Cumpston, T. Li, M. J. Page, & V. A. Welch (Eds.), Cochrane Handbook for Systematic Reviews of Interventions. Cochrane

Delgado-Alonso, C., Díez-Cirarda, M., Pagán, J., Pérez-Izquierdo, C., Oliver-Mas, S., Fernández-Romero, L., Martínez-Petit, Á., Valles-Salgado, M., Gil-Moreno, M. J., Yus, M., Matías-Guiu, J., Ayala, J. L., & Matias-Guiu, J. A. (2025). Unraveling brain fog in post-COVID syndrome: Relationship between subjective cognitive complaints and cognitive function, fatigue, and neuropsychiatric symptoms. Eur J Neurol, 32(1), e16084. 10.1111/ene.16084

Delis, D. C., Kaplan, E., & Kramer, J. H. (2001). Delis-Kaplan executive function system. Psychological Corporation.

Delis, D. C., Kramer, J. H., Kaplan, E., & Ober, B. A. (1994). California verbal learning tests, children’s version manual. Psychological Corporation.

Di Toro, A., Bozzani, A., Tavazzi, G., Urtis, M., Giuliani, L., Pizzoccheri, R., Aliberti, F., Fergnani, V., & Arbustini, E. (2021). Long COVID: long-term effects? European Heart Journal Supplements, 23(Suppl E), E1–E5. 10.1093/eurheartj/suab080

Diamond, A. (2013). Executive functions. Annual Review of Psychology, 64, 135–168. 10.1146/annurev-psych-113011-143750

Ertesvag, N. U., Iversen, A., Blomberg, B., Ozgumus, T., Rijal, P., Fjelltveit, E. B., Cox, R. J., Langeland, N., & Bergen, C.-r. g. (2023). Post COVID-19 condition after delta infection and omicron reinfection in children and adolescents. EBioMedicine, 92, 104599. 10.1016/j.ebiom.2023.104599

Fanshawe, J. B., Sargent, B. F., Badenoch, J. B., Saini, A., Watson, C. J., Pokrovskaya, A., Aniwattanapong, D., Conti, I., Nye, C., Burchill, E., Hussain, Z. U., Said, K., Kuhoga, E., Tharmaratnam, K., Pendered, S., Mbwele, B., Taquet, M., Wood, G. K., Rogers, J. P.,…Leek, C. E. (2024). Cognitive domains affected post-COVID-19; a systematic review and meta-analysis. European Journal of Neurology, e16181. 10.1111/ene.16181

Fisher, A. V. (2019). Selective sustained attention: a developmental foundation for cognition. Current Opinion in Psychology, 29, 248–253. 10.1016/j.copsyc.2019.06.002

Frolli, A., Ricci, M. C., Di Carmine, F., Lombardi, A., Bosco, A., Saviano, E., & Franzese, L. (2021). The Impact of COVID-19 on Cognitive Development and Executive Functioning in Adolescents: A First Exploratory Investigation. Brain Sciences, 11(9). 10.3390/brainsci11091222

Gjerdevik, M., & Heuch, I. (2014). Improving the error rates of the Begg and Mazumdar test for publication bias in fixed effects meta-analysis. BMC Medical Research Methodology, 14(1), 109–109. 10.1186/1471-2288-14-109

Gonzalez Aleman, G., Vavougios, G. D., Tartaglia, C., Uvais, N. A., Guekht, A., Hosseini, A. A., Lo Re, V., Ferreccio, C., D’Avossa, G., Zamponi, H. P., Figueredo Aguiar, M., Yecora, A., Ul Haq Katshu, M. Z., Stavrou, V. T., Boutlas, S., Gourgoulianis, K. I., Botero, C., González Insúa, F., Perez-Lloret, S.,…de Erausquin, G. A. (2025). Age-dependent phenotypes of cognitive impairment as sequelae of SARS-CoV-2 infection [Original Research]. Frontiers in Aging Neuroscience, 16. 10.3389/fnagi.2024.1432357

Gonzalez-Aumatell, A., Bovo, M. V., Carreras-Abad, C., Cuso-Perez, S., Domenech Marsal, E., Coll-Fernandez, R., Goicoechea Calvo, A., Giralt-Lopez, M., Ensenat Cantallops, A., Moron-Lopez, S., Martinez-Picado, J., Sol Ventura, P., Rodrigo, C., & Mendez Hernandez, M. (2022). Social, Academic, and Health Status Impact of Long COVID on Children and Young People: An Observational, Descriptive, and Longitudinal Cohort Study. Children, 9(11). 10.3390/children9111677

Gugliotta, M., Bisiacchi, P. S., Cendron, M., Tressoldi, P. E., & Vio, C. (2009). BVN 12-18: Batteria per la Valutazione Neuropsicologica per L’Adolescenza. Erickson.

Guido, C. A., Lucidi, F., Midulla, F., Zicari, A. M., Bove, E., Avenoso, F., Amedeo, I., Mancino, E., Nenna, R., De Castro, G., Capponi, M., Cinicola, B. L., Brindisi, G., Grisoni, F., Murciano, M., Spalice, A., & Long-Covid Group of Department of Maternal, S. (2022). Neurological and psychological effects of long COVID in a young population: A cross-sectional study. Frontiers in Neurology, 13, 925144. 10.3389/fneur.2022.925144

Harvey, P. D. (2019). Domains of cognition and their assessment. Dialogues in Clinical Neuroscience, 21(3), 227–237. 10.31887/DCNS.2019.21.3/pharvey

Hedges, L. V. (1981). Distribution theory for Glass’s estimator of effect size and related estimators. Journal of Educational Statistics, 6(2), 107–128. 10.3102/10769986006002107

Hendry, A., Jones, E. J. H., & Charman, T. (2016). Executive function in the first three years of life: Precursors, predictors and patterns. Developmental Review, 42, 1–33. 10.1016/j.dr.2016.06.005

Higgins, J. P. T., Eldridge, S., & Li, T. (2019). Including variants on randomized trials. In J. P. T. Higgins, J. Thomas, J. Chandler, M. Cumpston, T. Li, M. J. Page, & V. A. Welch (Eds.), Cochrane Handbook for Systematic Reviews of Interventions. Cochrane. https://training.cochrane.org/handbook

Invernizzi, A., Renzetti, S., van Thriel, C., Rechtman, E., Patrono, A., Ambrosi, C., Mascaro, L., Cagna, G., Gasparotti, R., Reichenberg, A., Tang, C. Y., Lucchini, R. G., Wright, R. O., Placidi, D., & Horton, M. K. (2023). Covid-19 related cognitive, structural and functional brain changes among Italian adolescents and young adults: a multimodal longitudinal case-control study. medRxiv. 10.1101/2023.07.19.23292909

Jason, L. A., Islam, M., Conroy, K., Cotler, J., Torres, C., Johnson, M., & Mabie, B. (2021). COVID-19 Symptoms Over Time: Comparing Long-Haulers to ME/CFS. Fatigue, 9(2), 59–68. 10.1080/21641846.2021.1922140

Jason, L. A., Johnson, M., & Torres, C. (2023). Pediatric Post-Acute Sequelae of SARS-CoV-2 Infection. Fatigue, 11(2-4), 55–65. 10.1080/21641846.2022.2162764

Jason, L. A., & Sunnquist, M. (2018). The development of the DePaul Symptom Questionnaire: Original, expanded, brief, and pediatric versions. Frontiers in Pediatrics, 6, 330–330. 10.3389/fped.2018.00330

Jassat, W., Reyes, L. F., Munblit, D., Caoili, J., Bozza, F., Hashmi, M., Edelstein, M., Cohen, C., Alvarez-Moreno, C. A., & Cao, B. (2023). Long COVID in low-income and middle-income countries: the hidden public health crisis. Lancet, 402(10408), 1115–1117. 10.1016/S0140-6736(23)01685-9

Jirout, J., LoCasale-Crouch, J., Turnbull, K., Gu, Y., Cubides, M., Garzione, S., Evans, T. M., Weltman, A. L., & Kranz, S. (2019). How lifestyle factors affect cognitive and executive function and the ability to learn in children. Nutrients, 11(8). 10.3390/nu11081953

Joshi, S. H., Siddarth, P., & Lavretsky, H. (2024). Structural MRI correlates of cognitive and neuropsychiatric symptoms in Long COVID: a pilot study. Front Psychiatry, 15, 1412020. 10.3389/fpsyt.2024.1412020

Korkman, M., Kirk, U., & Kemp, S. (2007a). NEPSY-II second edition: Clinical and interpretive manual. The Psychological Corporation.

Korkman, M., Kirk, U., & Kemp, S. (2007b). NEPSY-II: A developmental neuropsychological assessment. The Psychological Corporation.

Kwan, A. T. H., Portnoff, J. S., Al-Kassimi, K., Singh, G., Hanafimosalman, M., Tesla, M., Gharibi, N., Ni, T., Guo, Z., Sonfack, D. J. N., Martyniuk, J., Arfaie, S., Mashayekhi, M. S., Mofatteh, M., Jeremian, R., Ho, K., Moscote-Salazar, L. R., Lee, A., Jawad, M. Y.,…McIntyre, R. S. (2024). Association of SARS-CoV-2 infection with neurological impairments in pediatric population: A systematic review. Journal of Psychiatric Research, 170, 90–110. 10.1016/j.jpsychires.2023.12.005

Lai, Y. J., Liu, S. H., Manachevakul, S., Lee, T. A., Kuo, C. T., & Bello, D. (2023). Biomarkers in long COVID-19: A systematic review. Frontiers in Medicine, 10, 1085988. 10.3389/fmed.2023.1085988

Lamontagne, S. J., Winters, M. F., Pizzagalli, D. A., & Olmstead, M. C. (2021). Post-acute sequelae of COVID-19: Evidence of mood & cognitive impairment. *Brain, Nehavior*, & Immunity, 17, 100347–100347. 10.1016/j.bbih.2021.100347

Levine, R. L. (2022). Addressing the Long-term Effects of COVID-19. JAMA, 328(9), 823–824. 10.1001/jama.2022.14089

Lewallen, S., & Courtright, P. (1998). Epidemiology in practice: Case-controlled studies. Community Eye Health, 11(28), 57–58. https://pmc.ncbi.nlm.nih.gov/articles/PMC1706071/pdf/jceh_11_28_057.pdf

Li, J., Zhou, Y., Ma, J., Zhang, Q., Shao, J., Liang, S., Yu, Y., Li, W., & Wang, C. (2023). The long-term health outcomes, pathophysiological mechanisms and multidisciplinary management of long COVID. Signal Transduction and Targeted Therapy, 8(1), 416. 10.1038/s41392-023-01640-z

Long COVID and kids: more research is urgently needed. (2022). Nature, 602(7896), 183. 10.1038/d41586-022-00334-w

Luedke, J. C., Vargas, G., Jashar, D. T., Malone, L. A., Morrow, A., & Ng, R. (2024). Neuropsychological functioning of pediatric patients with long COVID. Clinical Neuropsychologist, 38(8), 1855–1872. 10.1080/13854046.2024.2344455

Manly, T., Anderson, V., Nimmo-Smith, I., Turner, A., Watson, P., & Robertson, I. H. (2001). The differential assessment of children’s attention: the Test of Everyday Attention for Children (TEA-Ch), normative sample and ADHD performance. *Journal of Child Psychology*, Psychiatry, and Allied Disciplines, 42(8), 1065–1081. 10.1111/1469-7610.00806

Masserini, F., Nicotra, A., Forgione, A., Calcaterra, F., Perdixi, E., Di Vito, C., Carletti, A., Gallo, C., Doneddu, P. E., Terenghi, F., Pomati, S., Nobile-Orazio, E., Riva, A., Mavilio, D., & Pantoni, L. (2025). Operationalisation of post-COVID condition case definition in a comprehensive research protocol. Eur J Neurol, 32(1), e16543. 10.1111/ene.16543

Menezes, F., Palmeira, J. D. F., Oliveira, J. D. S., Arganaraz, G. A., Soares, C. R. J., Nobrega, O. T., Ribeiro, B. M., & Arganaraz, E. R. (2024). Unraveling the SARS-CoV-2 spike protein long-term effect on neuro-PASC. Front Cell Neurosci, 18, 1481963. 10.3389/fncel.2024.1481963

Migliavaca, C. B., Stein, C., Colpani, V., Barker, T. H., Ziegelmann, P. K., Munn, Z., Falavigna, M., & Prevalence Estimates Reviews-Systematic Review Methodology, G. (2022). Meta-analysis of prevalence: I(2) statistic and how to deal with heterogeneity. Research Synthesis Methods, 13(3), 363–367. 10.1002/jrsm.1547

Miller, C. M., Borre, C., Green, A., Funaro, M., Oliveira, C. R., & Iwasaki, A. (2024). Post-Acute sequelae of COVID-19 in pediatric patients within the United States: A Scoping Review. American Journal of Medicine, 100078. 10.1016/j.ajmo.2024.100078

Miskowiak, K. W., Pedersen, J. K., Gunnarsson, D. V., Roikjer, T. K., Podlekareva, D., Hansen, H., Dall, C. H., & Johnsen, S. (2023). Cognitive impairments among patients in a long-COVID clinic: Prevalence, pattern and relation to illness severity, work function and quality of life. Journal of Affective Disorders, 324, 162–169. 10.1016/j.jad.2022.12.122

Moola, S., Munn, Z., Tufanaru, C., Aromataris, E., Sears, K., Sfetcu, R., Currie, M., Qureshi, R., Mattis, P., Lisy, K., & Mu, P. F. (2017). Chapter 7: Systematic reviews of etiology and risk. In Joanna Briggs Institute Reviewer’s Manual. The Joanna Briggs Institute. https://reviewersmanual.joannabriggs.org/

Mukherjee, S., Singer, T., Venkatesh, A., Choudhury, N. A., Perez Giraldo, G. S., Jimenez, M., Miller, J., Lopez, M., Hanson, B. A., Bawa, A. P., Batra, A., Liotta, E. M., & Koralnik, I. J. (2025). Vaccination prior to SARS-CoV-2 infection does not affect the neurologic manifestations of long COVID. Brain Commun, 7(1), fcae448. 10.1093/braincomms/fcae448

Narayanan, S. N., Padiyath, S., Chandrababu, K., Raj, L., P, S. B., Ninan, G. A., Sivadasan, A., Jacobs, A. R., Li, Y. W., & Bhaskar, A. (2025). Neurological, psychological, psychosocial complications of long-COVID and their management. Neurol Sci, 46(1), 1–23. 10.1007/s10072-024-07854-5

Nasreddine, Z. S., Phillips, N. A., Bédirian, V., Charbonneau, S., Whitehead, V., Collin, I., Cummings, J. L., & Chertkow, H. (2005). The Montreal Cognitive Assessment, MoCA: A Brief Screening Tool For Mild Cognitive Impairment. Journal of the American Geriatrics Society, 53(4), 695–699. 10.1111/j.1532-5415.2005.53221.x

Ng, R., Vargas, G., Jashar, D. T., Morrow, A., & Malone, L. A. (2022). Neurocognitive and Psychosocial Characteristics of Pediatric Patients With Post-Acute/Long-COVID: A Retrospective Clinical Case Series. Arch Clin Neuropsychol, 37(8), 1633–1643. 10.1093/arclin/acac056

Oberauer, K. (2019). Working Memory and Attention - Response to Commentaries. Journal of Cognition, 2(1), 30. 10.5334/joc.79

Page, M. J., Higgins, J. P. T., & Sterne, J. A. C. (2024). Assessing risk of bias due to missing evidence in a meta-analysis. In J. P. T. Higgins, J. Thomas, J. Chandler, M. Cumpston, T. Li, M. J. Page, & V. A. Welch (Eds.), Cochrane Handbook for Systematic Reviews of Interventions. Cochrane.

Page, M. J., McKenzie, J. E., Bossuyt, P. M., Boutron, I., Hoffmann, T. C., Mulrow, C. D., Shamseer, L., Tetzlaff, J. M., Akl, E. A., Brennan, S. E., Chou, R., Glanville, J., Grimshaw, J. M., Hrobjartsson, A., Lalu, M. M., Li, T., Loder, E. W., Mayo-Wilson, E., McDonald, S.,…Moher, D. (2021). The PRISMA 2020 statement: an updated guideline for reporting systematic reviews. BMJ, 372, n71. 10.1136/bmj.n71

Patel, A. M. R., Gilpin, G., Koniotes, A., Warren, C., Xu, C., Burgess, P. W., & Chan, D. (2025). Clinic evaluation of cognitive impairment in post-COVID syndrome: Performance on legacy pen-and-paper and new digital cognitive tests. Brain Behav Immun Health, 43, 100917. 10.1016/j.bbih.2024.100917

Pazukhina, E., Andreeva, M., Spiridonova, E., Bobkova, P., Shikhaleva, A., El-Taravi, Y., Rumyantsev, M., Gamirova, A., Bairashevskaia, A., Petrova, P., Baimukhambetova, D., Pikuza, M., Abdeeva, E., Filippova, Y., Deunezhewa, S., Nekliudov, N., Bugaeva, P., Bulanov, N., Avdeev, S.,…Sechenov Stop, C. R. T. (2022). Prevalence and risk factors of post-COVID-19 condition in adults and children at 6 and 12 months after hospital discharge: a prospective, cohort study in Moscow (StopCOVID). BMC Medicine, 20(1), 244. 10.1186/s12916-022-02448-4

Petersen, M. S., Kristiansen, M. F., Hanusson, K. D., Danielsen, M. E., B, A. S., Gaini, S., Strom, M., & Weihe, P. (2021). Long COVID in the Faroe Islands: A Longitudinal Study Among Nonhospitalized Patients. Clinical Infectious Diseases, 73(11), e4058–e4063. 10.1093/cid/ciaa1792

Pickering, H. E., Peters, J. L., & Crewther, S. G. (2023). A Role for Visual Memory in Vocabulary Development: A Systematic Review and Meta-Analysis. Neuropsychological Review, 33(4), 803–833. 10.1007/s11065-022-09561-4

Pour Mohammadi, S., Etesamipour, R., Mercado Romero F., & Peláez, I. (2024). A Step Forward in Long COVID Research: Validating the Post-COVID Cognitive Impairment Scale. Eur J Investig Health Psychol Educ, 14(12), 3001–3018. 10.3390/ejihpe14120197

Rabiner, D. L., Carrig, M. M., & Dodge, K. A. (2016). Attention Problems and Academic Achievement: Do Persistent and Earlier-Emerging Problems Have More Adverse Long-Term Effects? Journal of Attention Disorders, 20(11), 946–957. 10.1177/1087054713507974

Roge, I., Smane, L., Kivite-Urtane, A., Pucuka, Z., Racko, I., Klavina, L., & Pavare, J. (2021). Comparison of Persistent Symptoms After COVID-19 and Other Non-SARS-CoV-2 Infections in Children. Frontiers in Pediatrics, 9, 752385. 10.3389/fped.2021.752385

Roman, A. S., Pisoni, D. B., & Kronenberger, W. G. (2014). Assessment of Working Memory Capacity in Preschool Children Using the Missing Scan Task. Infant Child Development, 23(6), 575–587. 10.1002/icd.1849

Rothbart, M. K., Sheese, B. E., Rueda, M. R., & Posner, M. I. (2011). Developing Mechanisms of Self-Regulation in Early Life. Emotion Review, 3(2), 207–213. 10.1177/1754073910387943

Rothstein, T. L. (2023). Cortical Grey matter volume depletion links to neurological sequelae in post COVID-19 “long haulers”. BMC Neurology, 23(1), 22. 10.1186/s12883-023-03049-1

Rucker, G., Carpenter, J. R., & Schwarzer, G. (2011). Detecting and adjusting for small-study effects in meta-analysis. Biometrical Journal, 53(2), 351–368. 10.1002/bimj.201000151

Rucker, G., Schwarzer, G., & Carpenter, J. (2008). Arcsine test for publication bias in meta-analyses with binary outcomes. Statistics in Medicine, 27(5), 746–763. 10.1002/sim.2971

Rucker, G., Schwarzer, G., Carpenter, J. R., & Schumacher, M. (2008). Undue reliance on I(2) in assessing heterogeneity may mislead. BMC Medical Reswarch Methodology, 8, 79. 10.1186/1471-2288-8-79

Ruff, H. A., & Rothbart, M. K. (2001). Attention in early development: Themes and variations. Oxford University Press.

Rumain, B., Schneiderman, M., & Geliebter, A. (2021). Prevalence of COVID-19 in adolescents and youth compared with older adults in states experiencing surges. PLoS One, 16(3), 1–9. 10.1371/journal.pone.0242587

Sachdev, P. S., Blacker, D., Blazer, D. G., Ganguli, M., Jeste, D. V., Paulsen, J. S., & Petersen, R. C. (2014). Classifying neurocognitive disorders: the DSM-5 approach. *Nature Reviews*, Neurology, 10(11), 634–642. 10.1038/nrneurol.2014.181

Scarselli, V., Calderoni, D., Terrinoni, A., Davico, C., Pruccoli, G., Denina, M., Carducci, C., Smarrazzo, A., Martucci, M., Presicce, M., Marcotulli, D., Arletti, L., Ferrara, M., Garazzino, S., Mariani, R., Campana, A., & Vitiello, B. (2023). A Neuropsychiatric Assessment of Children with Previous SARS-CoV-2 Infection. Journal of Clinical Medicine, 12(12). 10.3390/jcm12123917

Seessle, J., Waterboer, T., Hippchen, T., Simon, J., Kirchner, M., Lim, A., Muller, B., & Merle, U. (2022). Persistent Symptoms in Adult Patients 1 Year After Coronavirus Disease 2019 (COVID-19): A Prospective Cohort Study. Clinical Infectious Diseases, 74(7), 1191–1198. 10.1093/cid/ciab611

Sekendiz, Z., Morozova, O., Carr, M. A., Fontana, A., Mehta, N., Ali, A., Jiang, E., Babalola, T., Clouston, S. A. P., & Luft, B. J. (2024). Characterization of Change in Cognition Before and After COVID-19 Infection in Essential Workers at Midlife. Am J Med Open, 12, 100076. 10.1016/j.ajmo.2024.100076

Sherman, E., & Brooks, B. L. (2015). Child and adolescent memory profile (ChAMP). Psychological Assessment Resources.

Siegel, A. L. M., & Castel, A. D. (2018). The role of attention in remembering important item-location associations. Memory & Cognition, 46(8), 1248–1262. 10.3758/s13421-018-0834-4

Smith, A. (1983). Symbol digit modalities test. Western Psychological Services.

Squire, L. R. (1987). Memory and brain. Oxford University Press.

Sterky, E., Olsson-Akefeldt, S., Hertting, O., Herlenius, E., Alfven, T., Ryd Rinder, M., Rhedin, S., & Hildenwall, H. (2021). Persistent symptoms in Swedish children after hospitalisation due to COVID-19. Acta Paediatrica, 110(9), 2578–2580. 10.1111/apa.15999

Su, H., Yang, P. L., Eaton, T. L., Valley, T. S., Langa, K. M., Ely, E. W., & Thompson, H. J. (2024). Cognition, function, and mood post-COVID-19: Comparative analysis using the health and retirement study. PLoS One, 19(12), e0315425. 10.1371/journal.pone.0315425

Sudre, C. H., Murray, B., Varsavsky, T., Graham, M. S., Penfold, R. S., Bowyer, R. C., Pujol, J. C., Klaser, K., Antonelli, M., Canas, L. S., Molteni, E., Modat, M., Jorge Cardoso, M., May, A., Ganesh, S., Davies, R., Nguyen, L. H., Drew, D. A., Astley, C. M.,…Steves, C. J. (2021). Attributes and predictors of long COVID. Nature Medicine, 27(4), 626–631. 10.1038/s41591-021-01292-y

Townsend, J. P., Hassler, H. B., Lamb, A. D., Sah, P., Alvarez Nishio, A., Nguyen, C., Tew, A. D., Galvani, A. P., & Dornburg, A. (2023). Seasonality of endemic COVID-19. mBio, 14(6), e0142623. 10.1128/mbio.01426-23

Tseng, M. H., & Chow, S. M. (2000). Perceptual-motor function of school-age children with slow handwriting speed. American Journal of Occupational Therapy, 54(1), 83–88. 10.5014/ajot.54.1.83

Tso, W. W. Y., Wang, Y., Fong, D. Y. T., Kwan, M. Y. W., Ip, P., Chan, J. F. W., Leung, L. K., Chan, J. Y. K., Tsao, S. S. L., Chau, C. S. K., Yip, K. M., Hui, K. Y., Duque, J. S. R., Lau, Y. L., & Lee, T. M. C. (2024). Development and validation of the Post-COVID Symptom Scale for Children/Youth (PCSS-C/Y). Eur J Pediatr, 184(1), 81. 10.1007/s00431-024-05913-9

Vinter, A., Paxton, S., Witt, A., & Perruchet, P. (2010). Implicit learning, development, and education. In J. Didier & E. Bigand (Eds.), Rethinking physical and rehabilitation medicine. Springer Science & Business Media.

Wang, C., Ramasamy, A., Verduzco-Gutierrez, M., Brode, W. M., & Melamed, E. (2023). Acute and post-acute sequelae of SARS-CoV-2 infection: a review of risk factors and social determinants. Virology Journal, 20(1), 124. 10.1186/s12985-023-02061-8

Watanabe, A., Iwagami, M., Yasuhara, J., Takagi, H., & Kuno, T. (2023). Protective effect of COVID-19 vaccination against long COVID syndrome: A systematic review and meta-analysis. Vaccine, 41(11), 1783–1790. 10.1016/j.vaccine.2023.02.008

Weakley, K. E., Schikler, A., Green, J. V., Blatt, D. B., Barton, S. M., Statler, V. A., Feygin, Y., & Marshall, G. S. (2023). Clinical Features and Follow-up of Referred Children and Young People With Long COVID. Pediatric Infectious Disease Journal, 42(12), 1093–1099. 10.1097/INF.0000000000004081

Wechsler, D. (2003). Wechsler Intelligence Scale for Children-fourth edition (WISC-IV). Psychological Corporation.

Wechsler, D. (2008). WAIS–IV: Administration and scoring manual. Psychological Corporation.

Wechsler, D. (2011). Wechsler abbreviated scale of intelligence–Second edition (WASI-II). Pearson Assessment.

Wechsler, D. (2014). Wechsler intelligence scale for children–fifth edition (WISC-V). Pearson Assessment.

Wiguna, T., Setyawati Wr, N., Kaligis, F., & Belfer, M. L. (2012). Learning Difficulties and Working Memory Deficits among Primary School Students in Jakarta, Indonesia. Clin Psychopharmacol Neurosci, 10(2), 105–109. 10.9758/cpn.2012.10.2.105

Wijeratne, T., & Crewther, S. (2020). Post-COVID 19 Neurological Syndrome (PCNS); a novel syndrome with challenges for the global neurology community. Journal of Neurological Sciences, 419, 117179. 10.1016/j.jns.2020.117179

World Health Organisation. (2022, December 7). *Post COVID-19 condition (Long COVID)*. https://www.who.int/europe/news-room/fact-sheets/item/post-covid-19-condition

World Health Organisation. (2023, February 16). *A clinical case definition for post COVID-19 condition in children and adolescents by expert consensus*. https://www.who.int/publications/i/item/WHO-2019-nCoV-Post-COVID-19-condition-CA-Clinical-case-definition-2023-1

World Health Organisation. (2024). WHO COVID-19 Dashboard. Retrieved September 18, 2024 from https://data.who.int/dashboards/covid19/cases

Wulf Hanson, S., Abbafati, C., Aerts, J. G., Al-Aly, Z., Ashbaugh, C., Ballouz, T., Blyuss, O., Bobkova, P., Bonsel, G., Borzakova, S., Buonsenso, D., Butnaru, D., Carter, A., Chu, H., De Rose, C., Diab, M. M., Ekbom, E., El Tantawi, M., Fomin, V.,…Vos, T. (2022). Estimated Global Proportions of Individuals With Persistent Fatigue, Cognitive, and Respiratory Symptom Clusters Following Symptomatic COVID-19 in 2020 and 2021. JAMA, 328(16), 1604–1615. 10.1001/jama.2022.18931

Zeraatkar, D., Ling, M., Kirsh, S., Jassal, T., Shahab, M., Movahed, H., Talukdar, J. R., Walch, A., Chakraborty, S., Turner, T., Turkstra, L., McIntyre, R. S., Izcovich, A., Mbuagbaw, L., Agoritsas, T., Flottorp, S. A., Garner, P., Pitre, T., Couban, R. J., & Busse, J. W. (2024). Interventions for the management of long covid (post-covid condition): living systematic review. BMJ, 387, e081318. 10.1136/bmj-2024-081318

Zhang, D., Zhang, B., Wu, Q., Zhou, T., Tong, J., Lu, Y., Chen, J., Wang, H., Chisolm, D. J., Jhaveri, R., Kenney, R. C., Rothman, R. L., Rao, S., Williams, D. A., Hornig, M., Wang, L., Morris, J. S., Forrest, C. B., & Chen, Y. (2025). Racial/ethnic differences in post-acute sequelae of SARS-CoV-2 in children and adolescents in the United States. Nat Commun, 16(1), 878. 10.1038/s41467-024-55273-z

Zheng, Y. B., Zeng, N., Yuan, K., Tian, S. S., Yang, Y. B., Gao, N., Chen, X., Zhang, A. Y., Kondratiuk, A. L., Shi, P. P., Zhang, F., Sun, J., Yue, J. L., Lin, X., Shi, L., Lalvani, A., Shi, J., Bao, Y. P., & Lu, L. (2023). Prevalence and risk factor for long COVID in children and adolescents: A meta-analysis and systematic review. Journal of Infection and Public Health, 16(5), 660–672. 10.1016/j.jiph.2023.03.005

